# Prophylactic and reactive vaccination strategies for healthcare workers against MERS-CoV

**DOI:** 10.1101/2022.04.06.22273497

**Authors:** Daniel J Laydon, Simon Cauchemez, Wes R Hinsley, Samir Bhatt, Neil M Ferguson

## Abstract

Several vaccines candidates are in development against Middle East respiratory syndrome–related coronavirus (MERS-CoV), which remains a major public health concern. Using individual-level data on the 2013-2014 Kingdom of Saudi Arabia epidemic, we employ counterfactual analysis on inferred transmission trees (“who-infected-whom”) to assess potential vaccine impact. We investigate the conditions under which prophylactic “proactive” campaigns would outperform “reactive” campaigns (i.e. vaccinating either before or in response to the next outbreak), focussing on healthcare workers. Spatial scale is crucial: if vaccinating healthcare workers in response to outbreaks at their hospital only, proactive campaigns perform better, unless efficacy has waned significantly. However, campaigns that react at regional or national level consistently outperform proactive campaigns. Measures targeting the animal reservoir reduce transmission linearly, albeit with wide uncertainty. Substantial reduction of MERS-CoV morbidity and mortality is possible when vaccinating healthcare workers, underlining the need for at-risk countries to stockpile vaccines when available.

## Introduction

The SARS-CoV-2 pandemic has demonstrated the threat posed by novel coronaviruses, the need for effective vaccines and the challenges in optimal vaccine deployment. First identified in the Kingdom of Saudi Arabia (KSA) in 2012 [1–3], Middle East respiratory syndrome–related coronavirus (MERS-CoV) remains a major public health concern, with 2585 laboratory confirmed cases and 931 deaths having been reported in 27 countries as of 16 March 2021 [4].

Human-to-human transmission occurs primarily in nosocomial settings but is otherwise relatively rare [5], and dromedary camels constitute the animal reservoir [6–8]. However, the threat of major outbreaks arising from human-to-human transmission should not be underestimated: a single introduction of the virus to South Korea caused 186 cases and 38 deaths in May-July 2015 [9].

Like SARS-CoV-2, MERS-CoV infection can be asymptomatic or fatal [10]. MERS-CoV has a lower transmissibility but higher lethality than SARS-CoV-2, with the Infection Fatality Ratio (IFR) increasing broadly with age in each [10, 11]. While it is not known whether MERS-CoV can transmit presymptomatically or asymptomatically like SARS-CoV-2 [12], sporadic infections in people with no known animal contact or human exposures suggests that asymptomatic or sub-clinical transmission is possible [13]. Estimates of the MERS-CoV IFR vary widely by country and study: in KSA and the Middle East, estimates range from 22% to 69%, whereas in South Korea mortality estimates ranged between 15% and 48% [1]. The IFR calculated using only laboratory confirmed cases is to date 36%. Similarly, the reproduction number (*R*) is thought to have been below 1 in Saudi Arabia and the Middle East but between 2.5 and 8.1 in South Korea [1].

No antivirals or vaccines against MERS-CoV are yet licensed [14], although several vaccine candidates are in development [15–18]. The most advanced candidates are the University of Oxford & Janssen Vaccines ChAdOx1 MERS recombinant viral vector vaccine [14], which phase 1b trials have recently shown to be safe, and to elicit both antibody and T cell immune responses in humans [19], and Inovio Pharmaceuticals’ INO-4700 nucleic acid vaccine, which is currently in phase 2 trials [20, 21].

In this study, we build on previous work [6] that inferred transmission trees (“who infected whom” analysis) to estimate the MERS-CoV reproduction number, serial interval, and the contributions of the animal reservoir and human-to-human transmission to the 2013-2014 epidemic in the KSA. This dataset is a detailed individual-level line list of a well described epidemic, and so allows us to address several questions: what would be the impact of a viable vaccine? Is it possible to substantially reduce morbidity and mortality if vaccinating healthcare workers only? What would be the optimal strategy to limit case numbers and deaths, and would this optimal strategy depend upon either vaccine efficacy or its duration of protection? If a vaccine cannot be assumed to maintain its efficacy permanently, would a “reactive” vaccination campaign, whereby vaccinations start in response to an outbreak, reduce case numbers more than a prophylactic “proactive” campaign that may occur years before the next outbreak? To what extent would control measures targeting the animal reservoir mitigate an epidemic?

We further use the inferred transmission trees because they preserve spatial properties of the epidemic by estimating the contributions of nosocomial, within-region, between-region and animal reservoir transmission. Using this approach further limits model assumptions, in that downstream effects of vaccination can simply be recorded.

We focus our analysis on healthcare workers since nosocomial transmission of MERS-CoV renders them the most at-risk population, and because they could feasibly be identified and vaccinated, either in preparation of future outbreaks, or at pace in response to an ongoing outbreak.

## Methods

### Data

We employ a line list [6] that includes the day of symptom onset, hospital, region, sex, healthcare worker status and clinical outcome of 681 cases of the 2013-2014 MERS-CoV outbreak in the KSA. Cases with symptom onset between 1^st^ January 2013 and 31^st^ July 2014 were included in the analysis. These data were previously used [6] to probabilistically infer transmission trees within a Bayesian framework. In this study we employ a counterfactual analysis whereby we “prune” the inferred transmission trees to evaluate a given strategy, considering what would have happened if a MERS-CoV vaccine had been available to healthcare workers. We can therefore limit assumptions on the extent to which vaccination would prevent transmission, and instead simply delete cases and their secondary cases (in this instance nodes and branches of the transmission tree) and then record the outcome.

### Transmission tree inference

The transmission tree inference we use has been described previously [6], but is included in the supplementary information for completeness. Broadly, inference is in two parts: i) inference of parameters; and ii) data augmentation for the infector of each case. Joint posterior estimates of the parameters and augmented data are generated using Markov chain Monte Carlo (MCMC) sampling. Each posterior sample therefore includes, in addition to parameters associated with the reproduction number and serial interval, a transmission tree. The following section discusses the generation of counterfactual posterior samples for given values of vaccine efficacy and duration, using a variety of vaccination strategies.

### Counterfactual transmission trees with vaccination

If case *k* ∈ 𝕅 has infector *i(k)* ∈ 𝕅, and symptom onset at time *t_k_*∈ 𝕅, then let *ψ_k_* ⊂ 𝕅 denote the set of all secondary cases from case *k,* i.e.

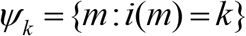

and so therefore (the infector of the secondary cases from case *k* is case *k*)

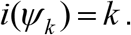

Then let 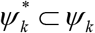 denote the set of secondary cases from case *k,* if case *k* had been vaccinated and protected. We assume that protection from disease also protects against onward transmission entirely, i.e. 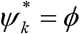, the empty set. Therefore, all downstream cases are deleted.

We consider two broad categories of vaccine campaign, which we name “proactive” and “reactive”. Under a proactive campaign, target vaccinees are vaccinated before an outbreak occurs, and so vaccinees are afforded at least some protection at the start of an outbreak. However, a proactive campaign has the obvious disadvantage that it is impossible to predict when the next outbreak will occur. Further, if the vaccine does not maintain its efficacy permanently, then efficacy may have waned substantially by the time the vaccine is actually needed. Recent experience with SARS-CoV-2 vaccines [22–27] suggests that waning of vaccine-induced immunity is highly plausible for a MERS-CoV vaccine.

Therefore, we also consider reactive vaccination campaigns that do not attempt to inoculate the population before the next outbreak, but rather in response to the current outbreak. While this has the disadvantage that the first cases in any outbreak will be left unprotected, it is possible that cases and deaths arising from slow react times would be outweighed by those arising from the suboptimal protection from a waning vaccine. Alternatively, elimination of downstream cases may mean that stopping as many early cases as possible, even with a vaccine of diminished efficacy, is more important than having the highest efficacy possible for the majority of would-be cases.

An advantage of a reactive campaign is that the vaccine would provide its maximum protection even if waning is substantial. However, the success of such a campaign is dependent on the speed with which target vaccinees are vaccinated, and the delay between vaccination and protection. We consider three levels of reactive campaign, whereby: i) cases within a given hospital would have been vaccinated in response to the first case in that hospital; ii) cases within a given region would have been vaccinated in response to the first case in that region (regardless the hospital in which the first case occurred); iii) cases within all regions would have been vaccinated in response to the first case in the country (regardless of the region in which the first case occurred). We are keen that each strategy we consider could at least in principle be implemented as policy, even if the required react times would prove challenging in practice.

Let the initial vaccine efficacy be denoted *VE*. In our main analysis, we assume exponential waning of efficacy, and if *D* is the vaccine’s mean efficacy duration, then efficacy after *t\** years post vaccination is given by

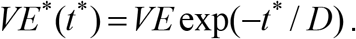

As a sensitivity analysis, we also consider slower waning of immunity using the sigmoidal Hill function, and so in this case

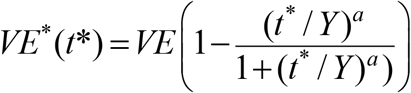

where *Y* is the efficacy half-life and *a* governs the speed of decline. We set *a* = 4 as a balance between allowing the vaccine to maintain its efficacy for longer than exponential waning, while also having a reasonably gradual decline [Figure 1].

**Figure 1:**
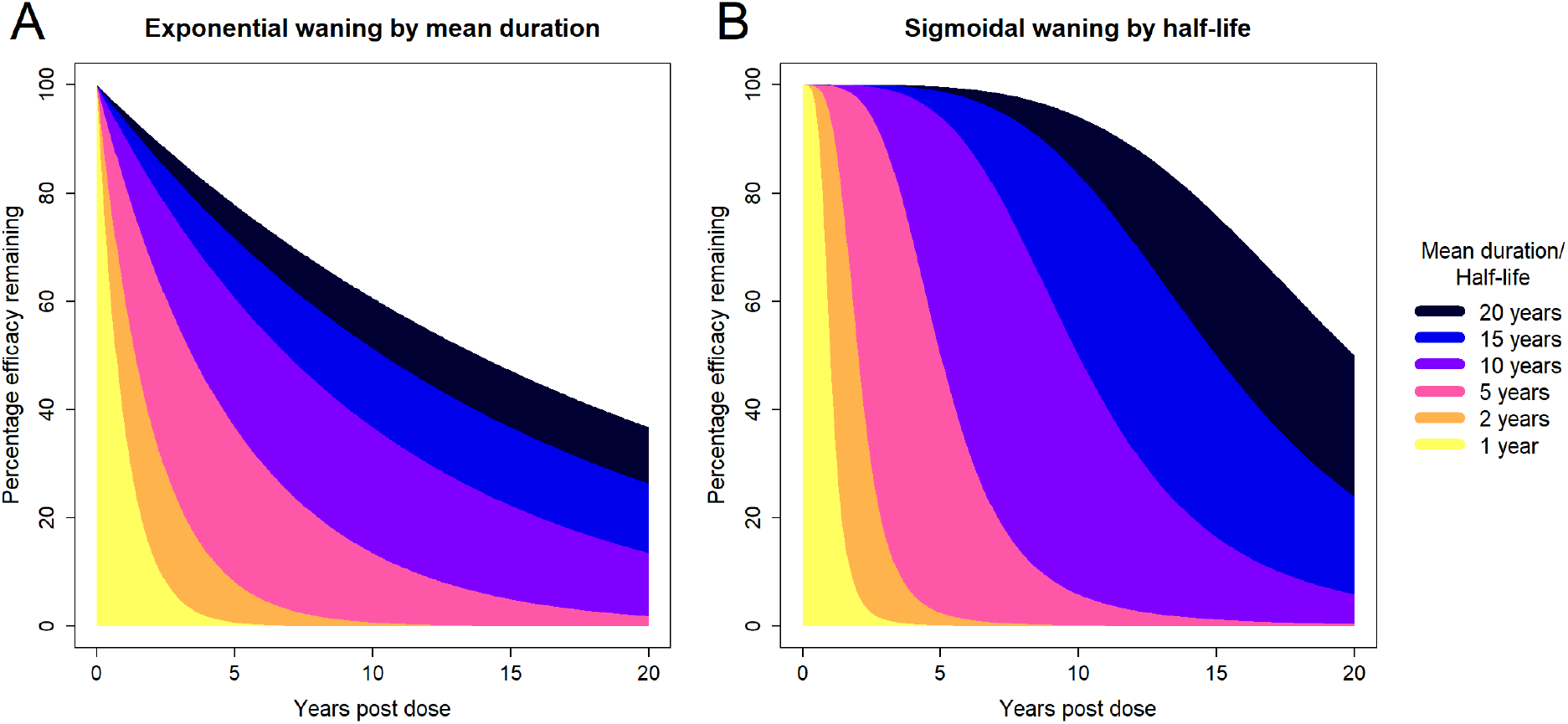
Percentage of initial vaccine efficacy remaining over time **A** by mean duration, assuming exponential waning; **B** by half-life, assuming sigmoidal waning. Mean durations and half-lives of 1, 2, 5, 10, 15 and 20 years are shown.

We consider mean durations (or half-lives if waning is sigmoidal) of 1, 2, 5, 10, 15, and 20 years, as well as no vaccine waning. We simulate values of 6 months, and 1 to 10 years for the time between vaccination and the next outbreak (which we term the “wait” for brevity).

If *S* denotes the set of people to be vaccinated, and *P_c_* is the coverage achieved in a campaign, then under a proactive strategy the probability *P_v,k_* that a given case *k* will be vaccinated is given by

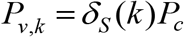

where *δ_S_* (*k*) is defined as

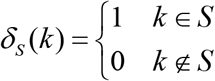

and the probability that case *k* will be protected, and deleted from the transmission tree, is

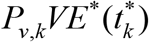

where *t_k_\** is the time post vaccination for case *k*. In this work, *S* is the set of healthcare workers, and we assume full coverage (i.e. *P_c_* = 1) of this group, although the effects of reduced coverage can simply be obtained through scaling. For example, 45% efficacy with 100% coverage is equivalent to 90% efficacy with 50% coverage.

Under a reactive campaign, a delay must be incorporated to account for first the react time *τ_I_* between the first case in a hospital (or region or country), and second the lag *τ_P_* between vaccination and protection. Hence the probability that case *k* will be vaccinated is given by the following product

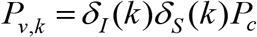

where *δ_I_* (*k*) is defined as

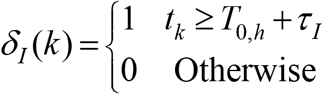

If a reactive campaign takes place at the level of a hospital, we define *T_0,h_* as the time of symptom onset of the first case in hospital *h*. However, if reacting at regional or national level, *T_0,h_* is defined as the time of symptom onset of the first case in region *h* or the entire country.

For reactive campaigns, only the vaccine’s initial efficacy is relevant (and not its duration of protection) as vaccine waning will be negligible in the time frames we consider between an outbreak and implementation. Here then the probability that case *k* will be protected and deleted from the transmission tree, is

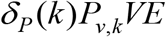

where *δ_P_* (*k*) is given by

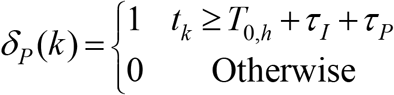

and where we vary *τ_I_* between 0 and 28 days in 2-day intervals. T cell responses to ChAdOx1 MERS peaked at 14 days, and while for antibodies the peak was observed at 28 days, antibody titres were still high at 14 days [19], and therefore we set *τ_P_* = 14 days. It is important to note that if the vaccine takes longer than 14 days-post-dose to confer protection, this is effectively already included in our analysis, as it is really the react time plus the time to protection that is important and so a longer time to protection is essentially a relabelling.

We investigate the effect of control measures aimed at limiting animal reservoir transmission, which we model as a simple proportion *γ* of reservoir infections that are stopped. That is, if in a particular posterior sample of a given model run, case *k* has *i(k) = 0,* then case *k* and all their downstream cases are deleted with probability *ς.* We consider camel control measure efficacies (values of *γ*) of 10%, 20%, 30%, 40% and 50%. Camel control measures are considered both in isolation and in combination with vaccination campaigns.

For a given strategy, we calculate the proportion of cases and deaths averted, and the change in transmission proportions (from e.g. the animal reservoir or within hospitals). The strategies we model are summarised in Table 1. In both proactive and reactive campaigns, we consider values of initial efficacy in 5% intervals between 5% and 100%.

**Table 1.** Modelled vaccination campaign strategies.

| Description<br>(Symbol) | Values simulated / Comments |
| --- | --- |
| Initial vaccine efficacy ($VE$) | Between 5% and 100% in intervals of 5% |
| Camel control measure effectiveness ($\zeta$) | 10%, 20%, 30%, 40%, 50% and 0% (i.e. no measures targeting animal reservoir), |
| **Proactive** |  |
| Mean duration ($D$) /<br>Half-life ($Y$) | 1, 2, 5, 10, 15, 20-year duration (or half-life if considering sigmoidal waning) and infinite duration (i.e. no vaccine waning) |
| Wait between vaccination and next outbreak ($t^*$) | 1 year to 10 years, and 6 months. |
| **Reactive** |  |
| Spatial reaction level for reactive campaign | i) hospital; ii) regional; iii) national |
| React time ($\tau_l$) | 0 to 28 days in 2-day intervals, between start of (hospital, regional or national) outbreak and vaccination |
| Time between vaccination and immunity ($\tau_p$) | 14 days (zero immunity assumed between 0 and 13 days post vaccination) |

### Model fitting

In the inference of transmission trees and parameter posteriors, all priors are uniform and fitted on a log scale. We performed 55,000 iterations with a burn-in period of 5,000, thinning every 5 iterations, resulting in 10,000 posterior samples per model run. Convergence is assessed visually. The model is coded in C++ and R version 4.0.5 [28], using packages “ggplot2” [29] and “igraph” [30] for plotting. All model code, (anonymised) data, and precompiled binaries are available at https://github.com/dlaydon/MERS_VacTrees.

## Results

### Data

In the 18 months from the start of 2013, there were 681 MERS cases, where date of symptom onset and patient’s hospital was reported, of which 534 (78%) were symptomatic at presentation and 276 (41%) were fatal. 187 (28%) of cases were in healthcare workers (HCWs), among whom there were 15 deaths, giving an 8% case-fatality ratio among healthcare workers, and comprising only 5% of all 276 deaths. The case-fatality ratio among non-HCWs was 53% [Figure 2].

**Figure 2:**
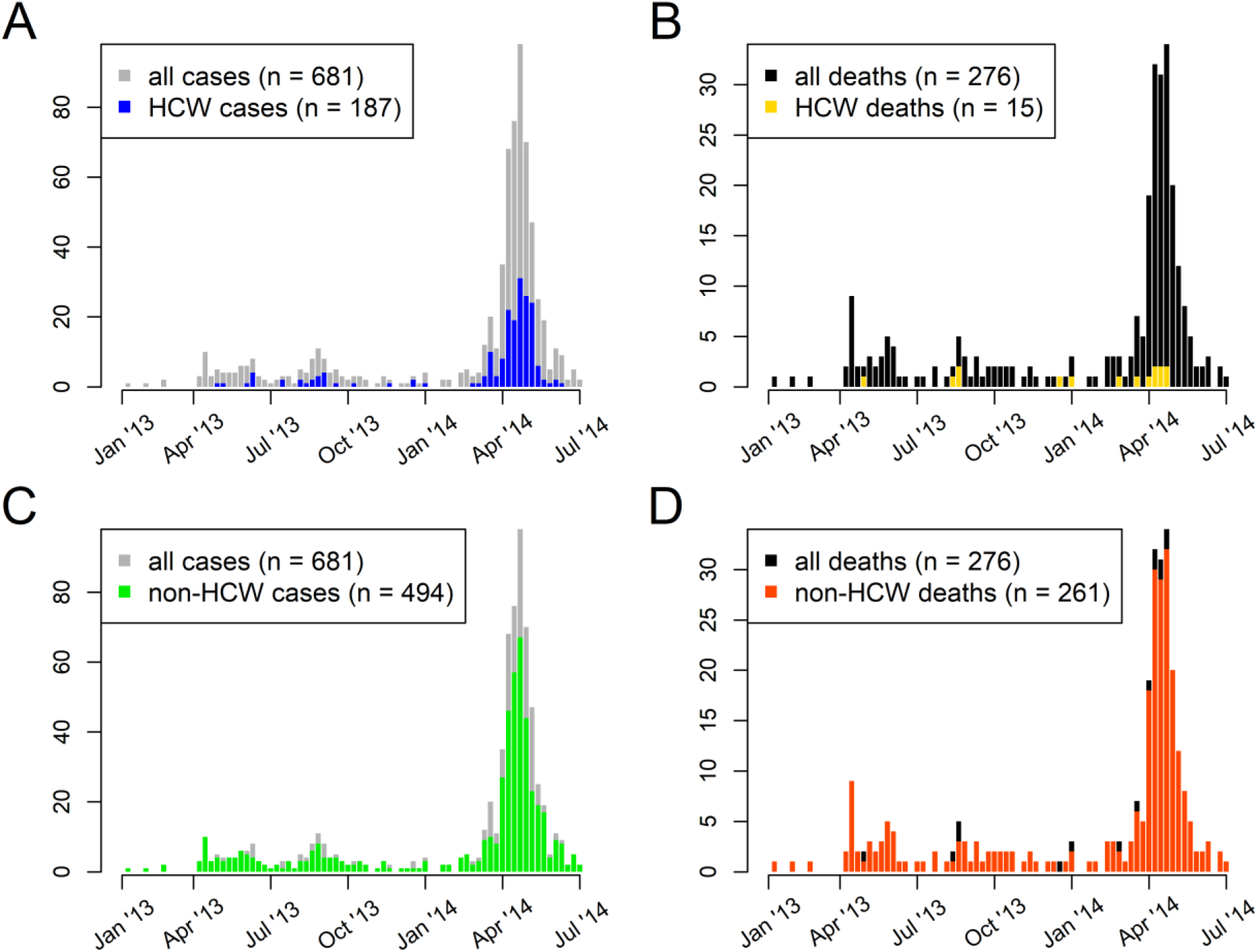
Weekly incidence of MERS-CoV cases (**A**, **C)** and deaths (**B**, **D**) for 2013-2014 outbreaks, among healthcare workers (HCW) (**A**, **B**) and non-healthcare workers (non-HCW) (**C**, **D**).

### Example model output

Figure 3 shows a series of transmission trees (“who-infected-whom” plots) from example model runs with and without vaccination. The trees show the contribution to transmission from the animal reservoir, as well as transmission within hospitals, between hospitals but within regions, and between regions.

**Figure 3:**
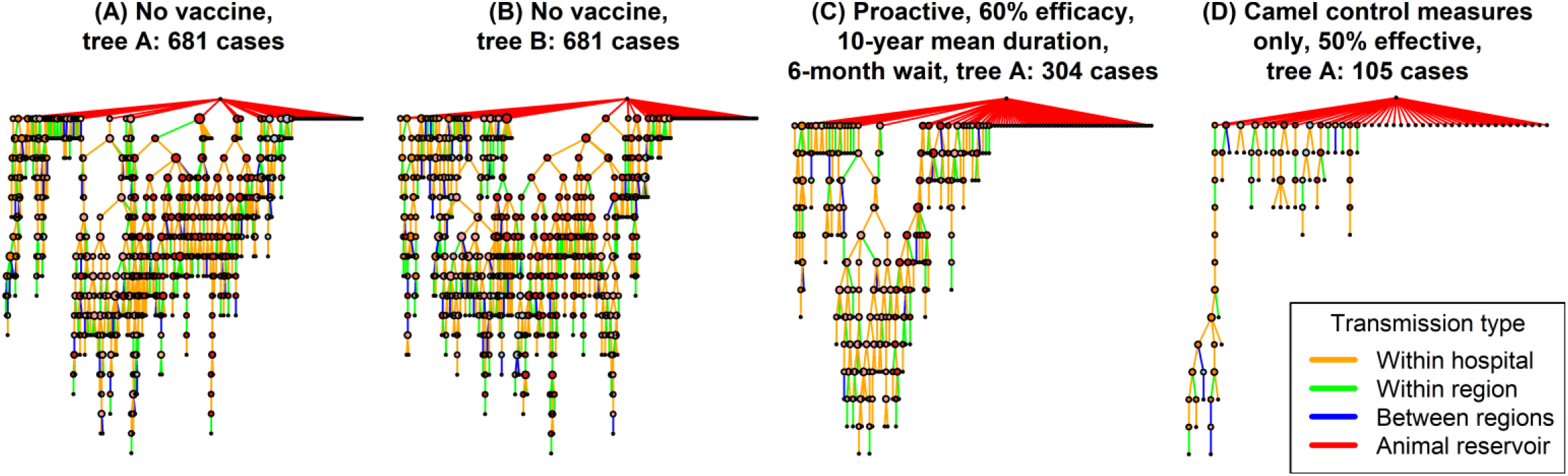
Example transmission trees. **(A)** No vaccine, inferred tree A. **(B)** No vaccine, inferred tree B. **(C)** 60% efficacious vaccine with 10-year mean duration, given in proactive campaign that occurs 6 months before the next outbreak. (**D)** Only control measures targeting transmission from the animal reservoir are considered, 50% efficacy assumed.

### Interventions targeting camels only

If humans are unvaccinated and interventions solely target the animal reservoir [Figure S1], then the mean proportions of cases and deaths averted is largely identical to the effectiveness of control measures. However, credible intervals are very wide, reflecting the unequal contribution of early cases in each local outbreak to the overall epidemic.

### Proactive campaigns

Under a proactive strategy, healthcare workers are vaccinated in anticipation of the next outbreak, and therefore all vaccinees have at least some protection from its outset. However, a proactive strategy depends on the extent of vaccine waning. Therefore success is a function of initial efficacy, duration and the wait time until the next outbreak, where the latter cannot be known in advance.

In the absence of camel control measures, under an optimistic scenario of 90% efficacy with a 20-year mean duration and only 6 months until the next outbreak, 64% (95% CrI: 54% - 74%) of cases and 51% (95% CrI: 39% - 64%) of deaths would be averted. However, if the next outbreak occurred 8 years after vaccination, then only 54% (95% CrI: 41% - 67%) of cases and 41% (95% CrI: 28% - 58%) of deaths would be averted. The 2013-2014 KSA MERS-CoV outbreak has been the only one of its scale [31], which suggests that the wait until next large outbreak (i.e. an outbreak that will most require vaccination) will be long.

Figure 4 shows the proportion of cases averted as a function of efficacy, duration, wait between vaccination and the next outbreak, and the effectiveness of camel control measures. Figure S2 shows the equivalent plots for the proportion of deaths averted. The success of a proactive campaign increases with the vaccine’s efficacy and duration of protection, and decreases with the wait until next outbreak. The wait between vaccination and outbreak is largely irrelevant if duration is long (e.g. 20 years), whereas the duration matters far less for short waits, and so here success is a function primarily of efficacy. In any case, low efficacies (e.g. ≤25%) struggle to make any impact, achieving at most a 31% (95% CrI: 18% - 51%) reduction in cases and 23% (95% CrI: 10% - 42%) reduction in deaths. Short durations (≤ 2 years) similarly struggle unless waits are short (<1 year) and efficacy is at least moderate (e.g. ≥50%).

**Figure 4.**
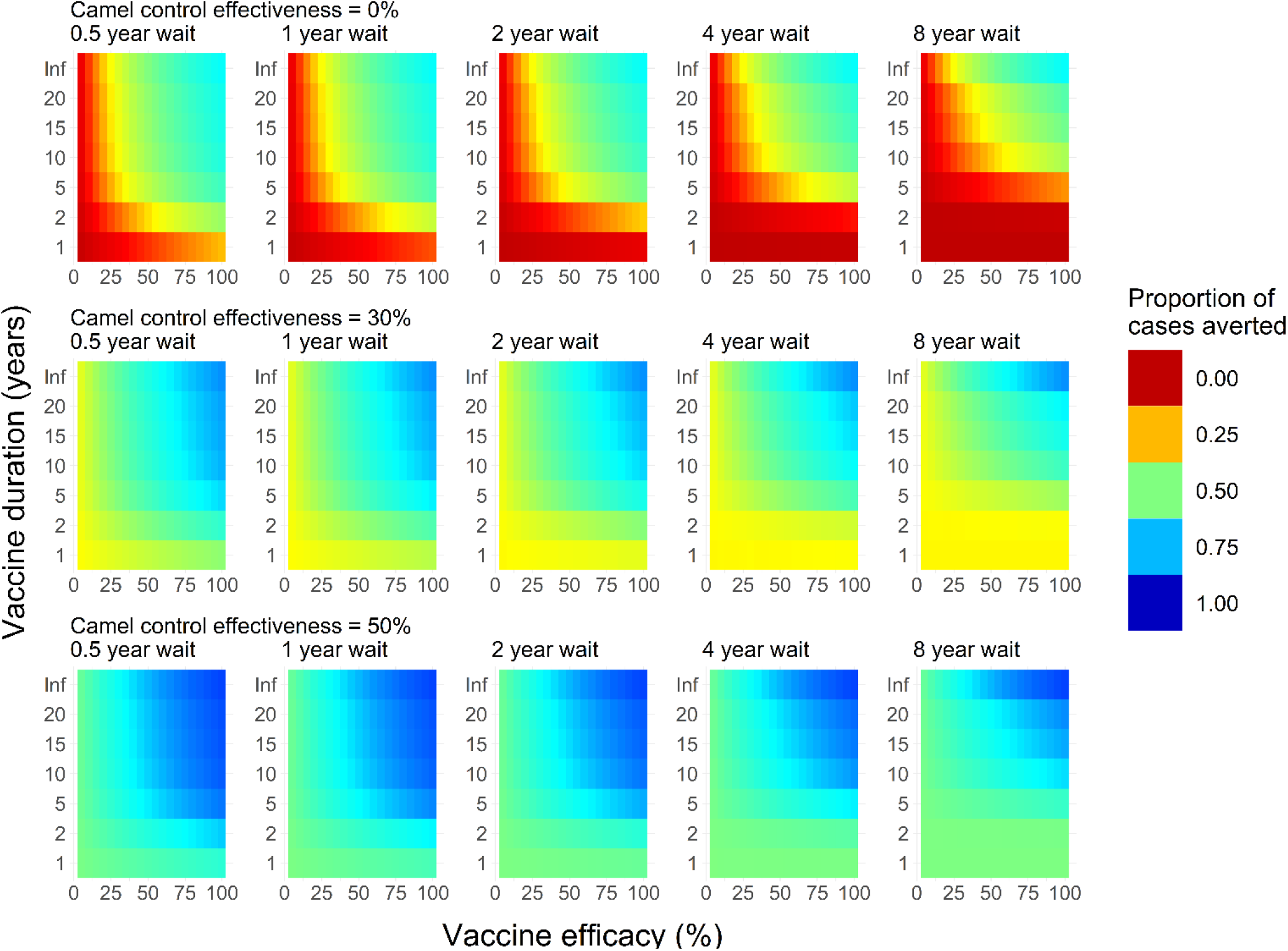
Each plot shows the mean posterior estimate of the proportion of cases averted by efficacy (x-axis) and mean duration (y-axis). Deeper blue colours indicate more averted cases. Each column of plots shows a given wait until the next outbreak after vaccination. Waits of 6 months, 1, 2, 4, and 8 years are shown. Each row of plots shows a given efficacy of camel control measures: values of 0% (i.e. no camel control measures), 10%, 30% and 50% are shown.

Adding measures targeting the animal reservoir can make large differences to proactive campaigns [Figure 4]. For example, 30% effective camel controls would improve the above optimistic scenario (90% efficacy, 20-year mean duration and 6 months wait) to 75% (95% CrI: 63% - 87%) of cases and 66% (95% CrI: 49% - 83%) of deaths. 50% effective camel control measures would improve this further to 82% (95% CrI: 69% - 92%) of cases and 75% (95% CrI: 57% - 90%) of deaths. Trends with efficacy, duration and wait time hold with the addition of camels, and the uncertainty in modelling camels in isolation is also present in combination with proactive campaigns. Lower credible intervals [Figure S3] show substantially less effective campaigns, whereas upper credible intervals [Figure S4] practically eliminate the epidemic for most values of efficacy, duration, wait time and camel control effectiveness that we considered.

### Reactive campaigns

In the likely event that vaccine efficacy wanes over time, even for a high efficacy and duration, a proactive campaign is still dependent on there being a sufficiently short wait until the next outbreak, at least in the absence of widespread and effective camel control measures.

Under a reactive campaign, an outbreak is already underway and so neither the wait until the next outbreak, nor the vaccine’s duration of protection are relevant. However, the react time between the first case of an outbreak and the implementation of a vaccination campaign will determine how a vaccine will fare. We model react times in 2-day intervals between 0 and 28 days and assume it takes 14 days for the vaccine to elicit an immune response.

In the ideal reactive scenario, with a 100% efficacious vaccine with instant implementation, vaccinating all healthcare workers in response to the first case at hospital level would avert 59% (95% CrI: 51% - 68%) of cases and 48% (95% CrI: 38% - 58%) of deaths. Since healthcare workers constituted only 28% of cases, this discrepancy illustrates the disproportionate effect of removing downstream cases [Figure 5].

**Figure 5:**
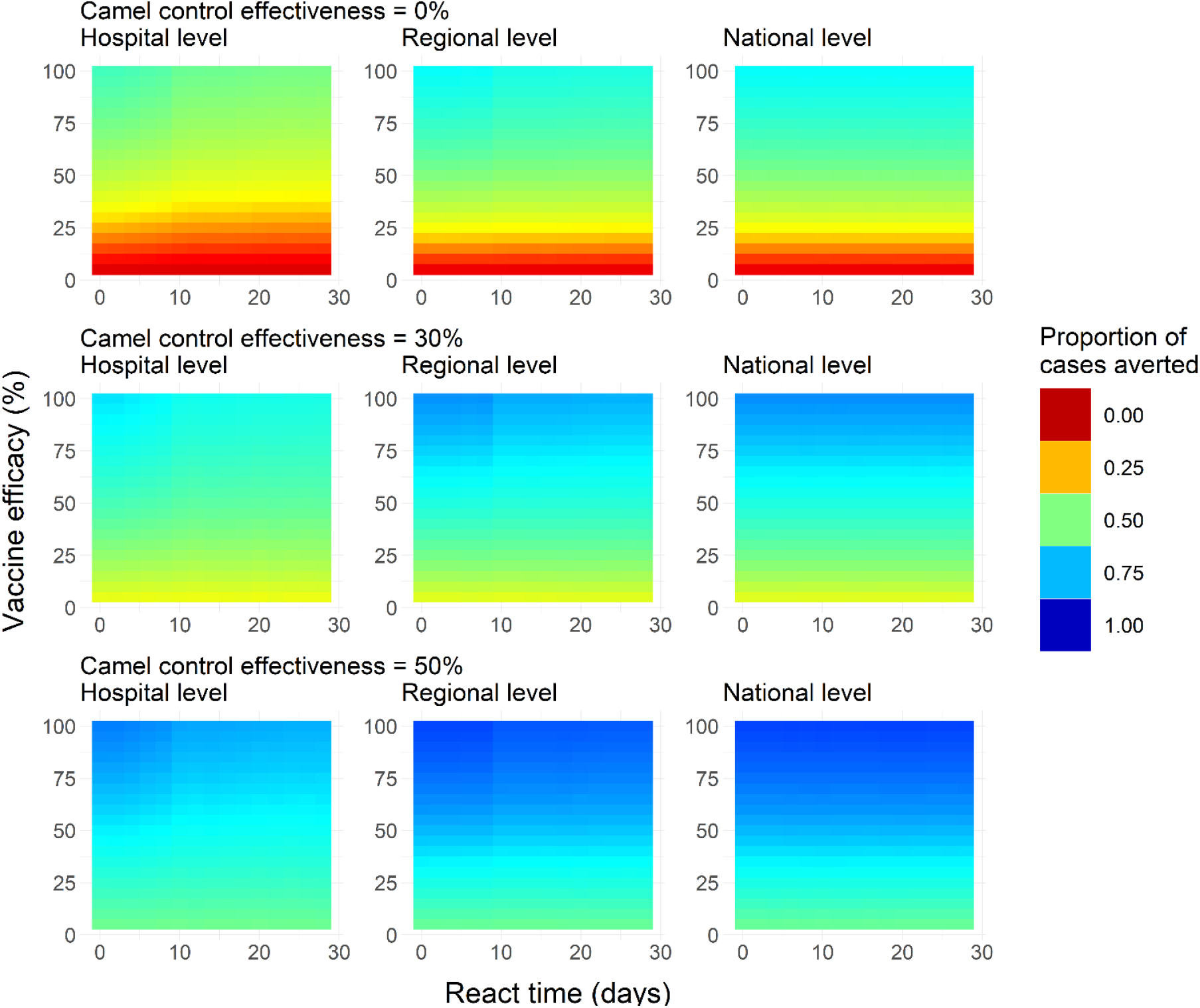
Mean posterior estimates for reactive campaigns of the proportion of cases averted, as a function of vaccine efficacy and react time. **Left**, **centre** and **right** columns show campaigns reactive at hospital, regional and national level. **Top, middle** and **bottom** rows show reactive campaigns in tandem with camel control measures of 0%, 30% and 50% respectively.

If a react time of 14 days is assumed, impact falls to 53% (95% CrI: 43% - 62%) of cases and 42% (95% CrI: 32% - 51%) of deaths, and for a 28-day react time falls further to 51% (95% CrI: 41% - 61%) of cases and 41% (95% CrI: 30% - 51%) of deaths. A vaccine efficacy of 50% with react times zero, 14 and 28 days would respectively reduce cases by 41% (95% CrI: 28% - 55%), 36% (95% CrI: 24% - 51%), and 35% (95% CrI: 22% - 50%) [Figure 5], and deaths by 32% (95% CrI: 18% - 46%), 28% (95% CrI: 16% - 42%), and 27% (95% CrI: 15% - 42%) [Figure S5]. For reactive campaigns at hospital level, react times beyond approximately 10 days are irrelevant, and success is a function of efficacy only. Lower and upper credible intervals are shown in Figures S6-S7, showing wide ranges in impact, as observed for proactive campaigns.

Greater impact can be achieved where a campaign reacts at regional level [Figure 5], where react times matter less than for hospital-level reactions. A perfect vaccine deployed instantaneously to healthcare workers would achieve a 69% (95% CrI: 61% - 77%) reduction in cases and a 55% (95% CrI: 44% - 67%) reduction in deaths. Assuming a 14-day react time, this falls to 66% (95% CrI: 58% - 74%) of cases and 51% (95% CrI: 41% - 62%) of deaths, and a 28-day react time reduces 65% (95% CrI: 58% - 73%) of cases and 50% (95% CrI: 41% - 62%) of deaths. Therefore, the react time makes less difference than at hospital level. Reacting at national level offers little further improvement [Figure 5], although interestingly the impact is the same regardless of whether the react time takes zero, 14 or 28 days, reducing cases by 69% (95% CrI: 61% - 77%) and deaths by 55% (95% CrI: 45% - 67%) in each instance.

The above national-level reductions are the best that can be achieved from reactive campaigns without camel control measures. For 30% effective camel controls, this maximum impact increases to 78% (95% CrI: 68% - 88%) of cases and 68% (95% CrI: 54% - 84%) of deaths averted, whereas with 50% effective camel controls, 84% (95% CrI: 74% - 93%) of cases and 78% (95% CrI: 62% - 91%) of deaths can be averted. Again, these results are insensitive to whether the national-level react time is zero days, 14 days, or 28 days.

The gains that can be achieved from reacting at regional or national level versus hospital level vary by efficacy and react time [Figure S8]. On average, across all efficacies and react times considered, regional and national level reactions offer a 31% and 34% improvement over hospital level reactions. The level at which a reactive campaign takes place is more important for longer react times. For example, there are greater differences (and ratios) between regional and hospital level reactions for an implementation that takes 28-day react time than for an instantaneous implementation. Broadly, the ratio of cases averted between reactive campaign levels decreases with efficacy, although this is mostly due to the limited impact of hospital-based reactive campaigns with low efficacy. However, the absolute difference increases with efficacy. At very long react times, national offers a slight improvement over regional and is approximately 5% better for efficacies above e.g. 60% [Figure S8]. Trends are largely the same for the number of deaths averted.

### Proactive vs. reactive campaigns

Figure 6 shows the ratios of cases averted between proactive and reactive campaigns. If vaccine efficacy does not wane, then the wait time until the next outbreak is irrelevant, and so proactive campaigns will always outperform reactive campaigns. Otherwise, a regional level reactive campaign with a 28-day react time is far superior to a proactive campaign, except where both vaccine duration is long (≥15 years) and the wait is short (≤1 year). National-level reactive campaigns were superior to proactive campaigns in every scenario we considered. Proactive campaigns require less stringent conditions to outperform hospital-level reactive campaigns, although even here reactive campaigns are superior in approximately half our simulations [Figure 6], and more often if the react time is reduced to say 8 days [Figure S9].

**Figure 6.**
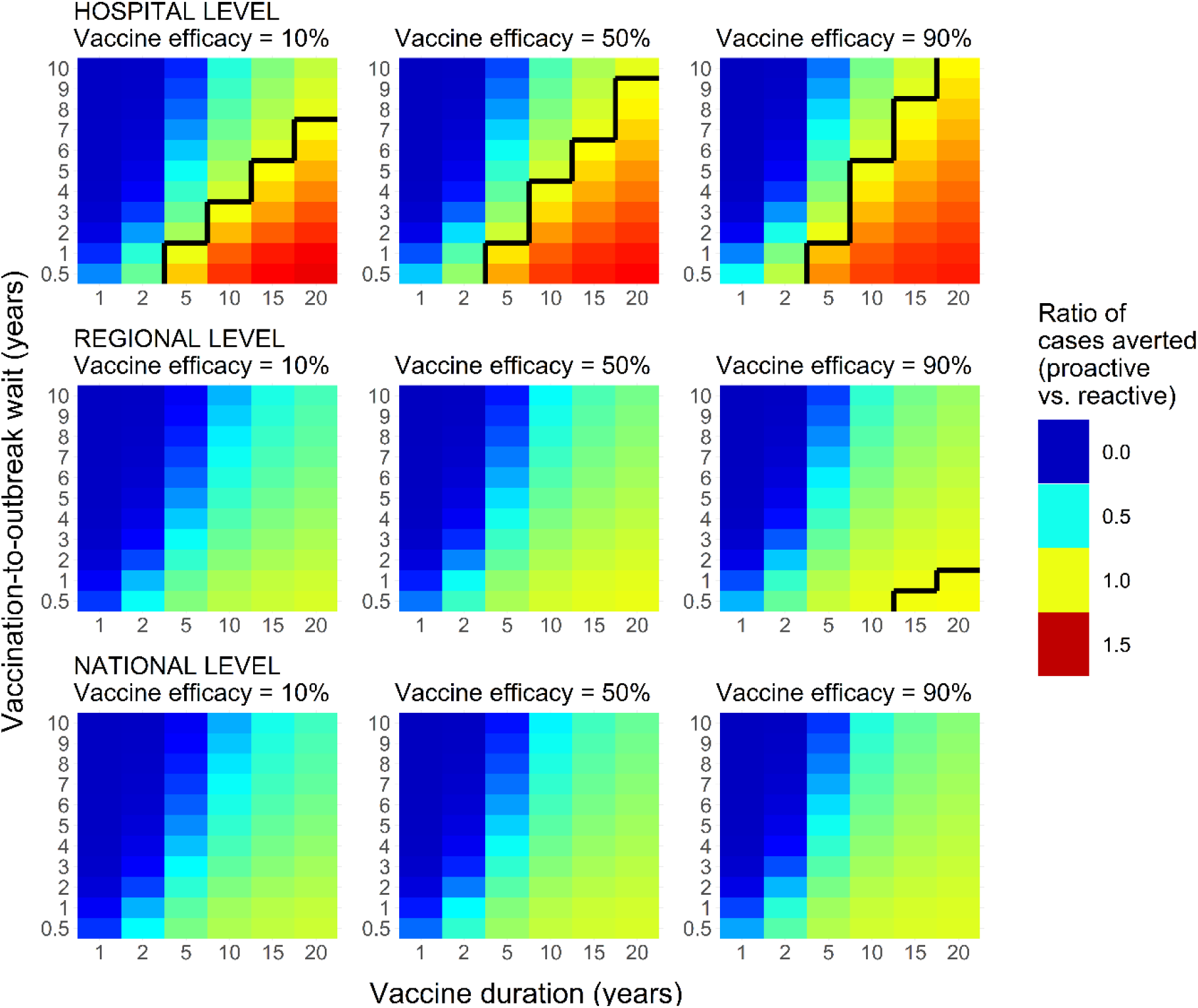
Each plot shows the ratio of cases averted between proactive and reactive (regional) campaigns (proactive / reactive), varying with duration of protection and wait until outbreak after vaccination. Proactive campaigns are compared to reactive campaigns at hospital level (top row), regional level (middle row) and national level (bottom row). Left to right columns show vaccine efficacies of 10%, 50% and 90%. A react time of 28 days is assumed in all plots, as well as a 14-day lag between vaccination and immunity. Ratios less than 1 (left of black contour line) indicate that a reactive campaign averts more cases.

The above trends hold almost irrespective of vaccine efficacy. Trends are also relatively insensitive to camel control measure effectiveness [Figure S10], although as their effectiveness increases, there is less proportional difference between proactive and reactive campaigns. Trends further hold when considering the ratio of deaths averted [Figure S11], although hospital- and regional-level campaigns outperform proactive slightly less often.

There is an asymmetry in the relative performance of proactive and reactive campaigns (if the vaccine does not retain its efficacy permanently). Assuming 10% vaccine efficacy, proactive campaigns can at best avert 39% more cases than hospital-level reactive campaigns, or 31% more cases assuming 90% efficacy. However, regional and national reactive campaigns of a 10% efficacious vaccine can respectively avert approximately 97 and 100 times more cases than a proactive campaign, or 421 and 444 times more if vaccine efficacy of 90% is assumed, albeit with the important caveat than the very poor performance of proactive campaigns with short duration and long wait results in very small numbers being compared.

### Change in transmission type

Figures S12 and S13 respectively show the change in relative and absolute transmission contribution with vaccine efficacy for a number of example scenarios. Without human vaccination, interventions targeting only the animal reservoir simply reduce overall case numbers, and do not change the proportional contributions of each transmission type (nosocomial, regional, national and reservoir). Otherwise, the change in transmission proportions with efficacy is highly similar between strategies. Nosocomial transmission decreases with vaccine efficacy from approximately 60% to between approximately 45% and 50%, depending on the specific strategy. Transmission within hospital, region and nationally goes down and therefore relative contribution of animal reservoir goes up (approximately 12% of cases without vaccination rising to as much as 30% depending on the strategy) [Figure S12]. Trends in absolute case numbers also do not differ markedly between vaccination strategies [Figure S13], although regional reactive campaigns avert more nosocomial, regional and national transmission than hospital level reactive campaigns.

### Sensitivity analyses

We investigated the sensitivity of our results to the choice of vaccine waning model. Our main analysis considers waning of immunity to be exponential. However it may be that a slower decline with a sigmoidal function, would be more appropriate. We therefore reran our analysis of proactive campaigns using a Hill function [Figure 1], considering the vaccine’s half-life as opposed to its mean duration.

The relationships between the proportion of cases averted and efficacy, duration (in this instance half-life), and the wait until the next outbreak are largely the same as for our default exponential waning model [Figure S14], although longer half-lives (>5 years) perform slightly better and shorter half-lives (<5 years) perform slightly worse. In general though, the predicted impact of proactive campaigns is marginally greater when considering sigmoidal waning.

Assuming sigmoidal waning affects the relative performance of reactive and proactive campaigns [Figure S15]. Hospital- and regional-level reactive campaigns are more often eclipsed by proactive campaigns. However, nationally reactive campaigns are still superior to proactive campaigns in almost all simulations. Trends are again consistent across different values of vaccine efficacy, and with camel control effectiveness (not shown). Our results are reasonably robust to the choice of waning model.

We were further concerned that the timeframe under consideration could bias our comparison of reactive and proactive campaigns. If a campaign reacts to cases (whether at hospital, regional or national level) in 2013, then there is ample time to react before the majority of cases occur during 2014 [Figure 2]. Figures S16-S18 show the comparison between reactive and proactive campaigns after dividing the entire Jan 2013 – Jul 2014 dataset into three subsets (i) Jan-Jun 2013; ii) Jul-Dec 2013, and iii) Jan-Jul 2014). For all three subsets, any advantages previously enjoyed by hospital-level reactive campaigns against proactive suffer substantially, regardless of efficacy: proactive campaigns are usually better, except for short durations and long waits between vaccination and the next outbreak. Regional-level reactive campaigns are affected less consistently: they fare worse for each 2013 data subset, but are almost identical for the 2014 subset. However, campaigns that react at national level consistently outperform proactive campaigns, almost regardless of the timeframe considered, although there is little difference between proactive and reactive campaigns here.

It is important to note that where ratios are high, and therefore where proactive campaigns ostensibly avert many times the cases than reactive campaigns, there are very few cases to avert in the first place, particularly among healthcare workers [Figure 2]. Therefore, in situations where there are most cases where a vaccine is most needed, reactive policies at national level are best.

## Discussion

No vaccine against MERS-CoV has yet been licensed in humans. If and when such a vaccine becomes available, determining its optimal deployment is nontrivial. In this study, we analysed multiple vaccine campaign strategies as a function of efficacy and duration. Each strategy was evaluated by estimating multiple transmission trees (who-infected-whom), and then “pruning” them to generate counterfactual epidemics to determine the number of cases and deaths that a vaccine would prevent. Our intention is that all strategies considered could at least in principle be implemented, and therefore that our analysis will be relevant for policymakers. We considered vaccination of healthcare workers only, as they will be most easily vaccinated and most exposed, and therefore more cost-effective.

We considered the relative merits of proactive and reactive campaigns, where the fundamental difference between the two approaches is whether to vaccinate in anticipation of the next outbreak, or in response to the current outbreak. The success of a proactive campaign is a function of vaccine efficacy, duration and the wait time between vaccination and the next outbreak. Whereas for reactive campaigns, success is dependent on efficacy, react time and the spatial level at which a vaccination campaign reacts: in response to a hospital, regional or national outbreak. In all scenarios examined, vaccination has a greater effect on cases than deaths, likely because healthcare workers firstly constitute only 27% of cases, and secondly because they are younger than non-healthcare workers (mean age 39 vs. 51 years) and so are probably healthier.

Short durations or long waits (or both) greatly diminish the impact of a proactive campaign. While the wait time until the next major outbreak cannot be known in advance, still less its magnitude, given that the 2013-2014 KSA MERS-CoV outbreak was the only one of its scale [31], it is reasonable to think that the wait until the next large outbreak will be long. In contrast, we have not modelled reactive campaigns to depend on the vaccine’s duration, and so vaccinees are conferred maximum possible benefit, provided that vaccines can be administered and elicit an immune response before people would otherwise be infected.

Therefore, the spatial scale at which a campaign reacts is crucial. If each hospital reacts individually to its own outbreak, many cases (and their secondary cases) occur before vaccination or immunity. However, campaigns that react at regional or national level suffer far less from these delays and therefore can avert many more cases than their proactive equivalents, even where the time until the next outbreak is short and durations are long (although proactive campaigns are always better than reactive if vaccine efficacy does not wane). Interestingly, the relative performance of reactive and proactive campaigns does not depend on efficacy, and the introduction of measures to limit transmission from the animal reservoir does not affect the rank order of campaigns. Essentially, regionally and nationally reactive campaigns offer an opportunity to get ahead of the epidemic, and can be viewed as a proactive campaign but with a more certain and shorter wait time.

Our analysis is reasonably robust to whether vaccine waning is exponential or sigmoidal, but more sensitive to the choice of timeframe, in that hospital-level reactive campaigns are rarely superior to proactive. However, our main conclusions firstly that the spatial scale at which a vaccination campaign reacts is crucial, and secondly that nationally reactive campaigns are campaigns are the most effective way to reduce MERS-CoV case numbers and deaths, are strengthened. Further, focussing on the impact on a single smaller outbreak slightly misses the point: we are ultimately most interested in the maximum possible morbidity and mortality reductions over the widest possible timeframe. In effect this blurs the distinction between reactive and proactive campaigns: a reactive campaign against one outbreak can also be considered as a proactive campaign against a subsequent outbreak.

We are aware of some limitations in our analysis. We assume that all downstream cases of a successfully inoculated person are deleted, and this is unlikely to be completely true for two reasons. First, if downstream cases had not been infected by their index case, they may still have been infected through another route. Second, the vaccine may have differing efficacies against disease and transmission. A vaccine that inoculates against disease may not stop transmission or vice-versa. Additionally, we assume there is no age-dependency in vaccine efficacy, and recent experience with SARS-CoV-2 vaccines suggests that that some age-dependency is likely [32–34]. While properly accounting for these issues would improve our estimates, we do not have any data to inform such analysis, and so any attempt to do so would be overly speculative.

It is also possible that some sub-clinical infections were not detected, and are therefore missing from our line list. If such cases contributed meaningfully to transmission, then our results could be biased upwards, and arguably more so for reactive campaigns, as reactive campaigns might not react to index case in a hospital or region or country. We have also not accounted for any behavioural change or risk compensation in response to an available vaccine.

In considering camel control measures, we have assumed only a simple proportional reduction in contribution from animal reservoir, without specifying what this would entail (e.g. vaccination, better hygiene, or reduced physical contact between humans and camels), and clearly additional data to inform more precise analysis would be helpful.

To reduce the otherwise prohibitively large number of simulations, we have assumed no vaccine waning under reactive campaigns. However, unless duration was very short the effects of waning would be negligible, and in this instance, waning would still affect reactive campaigns less than proactive campaigns. On the other hand, we assume zero immunity until 14 days post vaccination, whereas in practice there would be at least some protection prior to this. Greater delays between vaccination and immunity would affect our results, but not the trends we describe, and would essentially constitute a relabelling (e.g. if the time was 10 or 18 days, the react times we list must be decreased or increased respectively by 4 days).

Because MERS-CoV outbreaks are relatively infrequent, traditional randomised controlled trials may not be feasible [35], and therefore vaccine efficacy or its wider effectiveness may be difficult to measure empirically, and this is even more so with the vaccine’s duration of protection. It is therefore useful to have an indication of the most effective strategies even if values of efficacy and duration are unknown. Unless the vaccine maintains its efficacy for a long time (>20 years) a reactive campaign at regional or national level will usually be superior to a proactive campaign.

Our analysis demonstrates that substantial reduction of MERS-CoV cases and deaths is possible even when vaccinating healthcare workers only, and underlines the need for countries at risk of MERS-CoV outbreaks to have reasonably large stockpiles of vaccines when they become available.

### Data sharing statement / Code availability

All model code, (anonymised) data, and precompiled binaries are available at https://github.com/dlaydon/MERS_VacTrees.

## Acknowledgements

DJL, WRH, SB and NMF acknowledge joint Centre funding from the UK Medical Research Council and Department for International Development (grant MR/R015600/1). DJL and NMF acknowledge funding from Vaccine Efficacy Evaluation for Priority Emerging Diseases (VEEPED) grant, (ref. NIHR: PR-OD-1017-20002) from the National Institute for Health Research. SB acknowledges The UK Research and Innovation (MR/V038109/1), the Academy of Medical Sciences Springboard Award (SBF004/1080), The BMGF (OPP1197730), Imperial College Healthcare NHS TrustBRC Funding (RDA02), The Novo Nordisk Young Investigator Award (NNF20OC0059309) and The NIHR Health Protection Research Unit in Modelling Methodology. SC acknowledges financial support from the Investissement d’Avenir program, the Laboratoire d’Excellence Integrative Biology of Emerging Infectious Diseases program (grant ANR-10-LABX-62-IBEID) and the INCEPTION project (PIA/ANR-16-CONV-0005). Views expressed do not necessarily represent those of the funders.

## Supplementary Methods

### Transmission tree inference

#### Parameter inference

The following has been presented previously [6], but is described here for ease of reference. MERS-CoV cases are partitioned into clusters, where we define a cluster as a group of cases from the same hospital, where the time lag between consecutive cases is at most 21 days. Let *ω_s_* denote the proportion of secondary cases with onset *s* days after the onset of their infector (i.e. the serial interval). *ω_s_* is assumed to be Gamma distributed and its mean and standard deviation are inferred from the data.

The MERS-CoV reproduction number is divided into three mutually exclusive parts: i) within-cluster; ii) between-cluster and within-region; iii) between-region. *R^c^_C_(m)* denotes the within-cluster reproduction number *R^c^_C_(0)* for cluster *c* after *m* cases, and its initial value *R^c^_C_(0)* is drawn from a gamma distribution with mean *R_c_* and standard deviation *σ_c_*. *R^c^_C_(m)* is given by *R^c^_C_(m) = R^c^_C_(0)(1+m)^-γ^*, where *γ* is a fitted parameter that describes the decline in the reproduction number with the number of cumulative cases, either due to infection control measures or to the depletion of susceptible individuals.

The mean number of animal reservoir infections *α_t_* on day *t* is given by *α_t_ = E_0_* exp*(αt)*, where *E_0_* is the initial number of reservoir infections at the beginning of the study period (i.e. January 1, 2013), and *α* is the (positive or negative) growth rate.

If case *k* ∈ 𝕅 has infector *i(k)* ∈ 𝕅, and is a member of cluster and region *c_k_* and *q_k_* respectively, then the individual reproduction numbers at within-cluster, between cluster and between-region level are given by

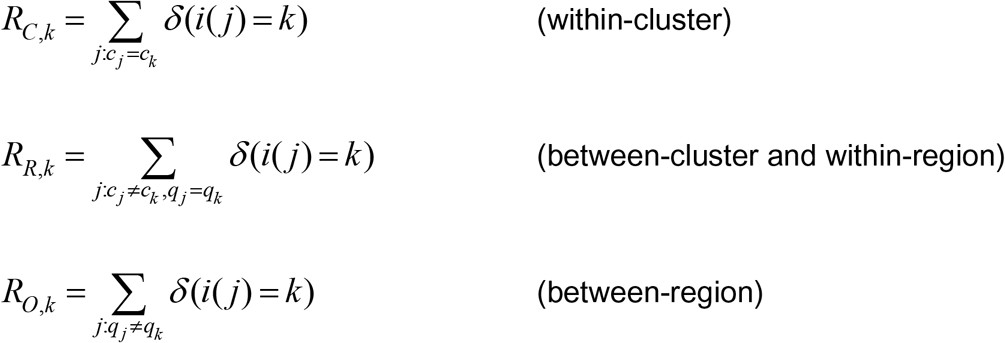

where *δ(i(j)=k)) =* 1 if *i(k) = j* and 0 otherwise. We set *i(k)* = 0 if the *k^th^* case was infected by the animal reservoir. Therefore the number of introductions *I_t_* on day *t* is given by 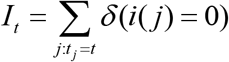.

The likelihood *L* is composed of three terms concerning: i) transmission between cases *L_k_^trans^*; ii) animal reservoir introduction *L_t_^intro^*; and iii) heterogeneity of transmission intensity between clusters *L^cluster^*.

*L_k_^trans^* is given by

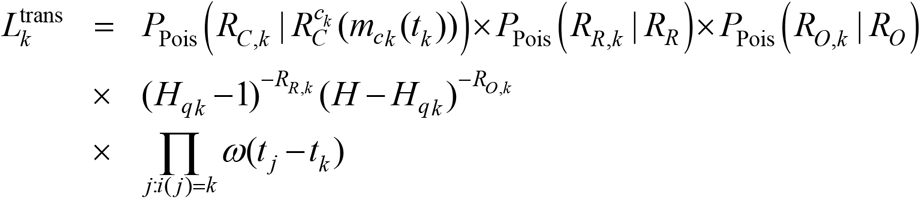

where *H* denotes the total number of hospitals in the data, and *H_q_* is the number of hospitals in region *q*. The first line of *L_k_^trans^* gives the probabilities of observing secondary cases arising from within-cluster, between-cluster and within-region, and between-region transmission. The second line of *L_k_^trans^* gives the probabilities that a secondary case outside of case *k*’s hospital will be in another hospital within region and outside region *q_k_*. The third line of *L_k_^trans^* gives the density of the serial interval.

The likelihood *L_t_^intro^* that there were *I_t_* infections from the animal reservoir on day *t* is

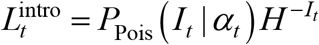

and the likelihood *L^cluster^* of the heterogeneity of transmission intensity is a product over clusters

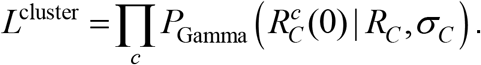

The full likelihood *L* is a product of the above terms

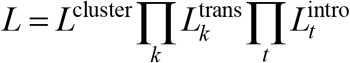

#### Data augmentation

In practice, the infector of each case is unknown, and so we use data augmentation and infer the joint posterior distribution of the parameters and infectors. For each possible source of infection *k* to case *j*, we define the following weights *w_k_*:

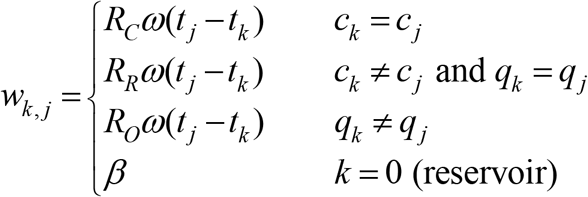

where *β* = 0.05. The above weights are normalised so that the probability *p_k,j_* that case *j* had infector *k* is given by

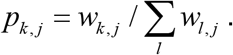

## Supplementary Figures

**Figure S1.**
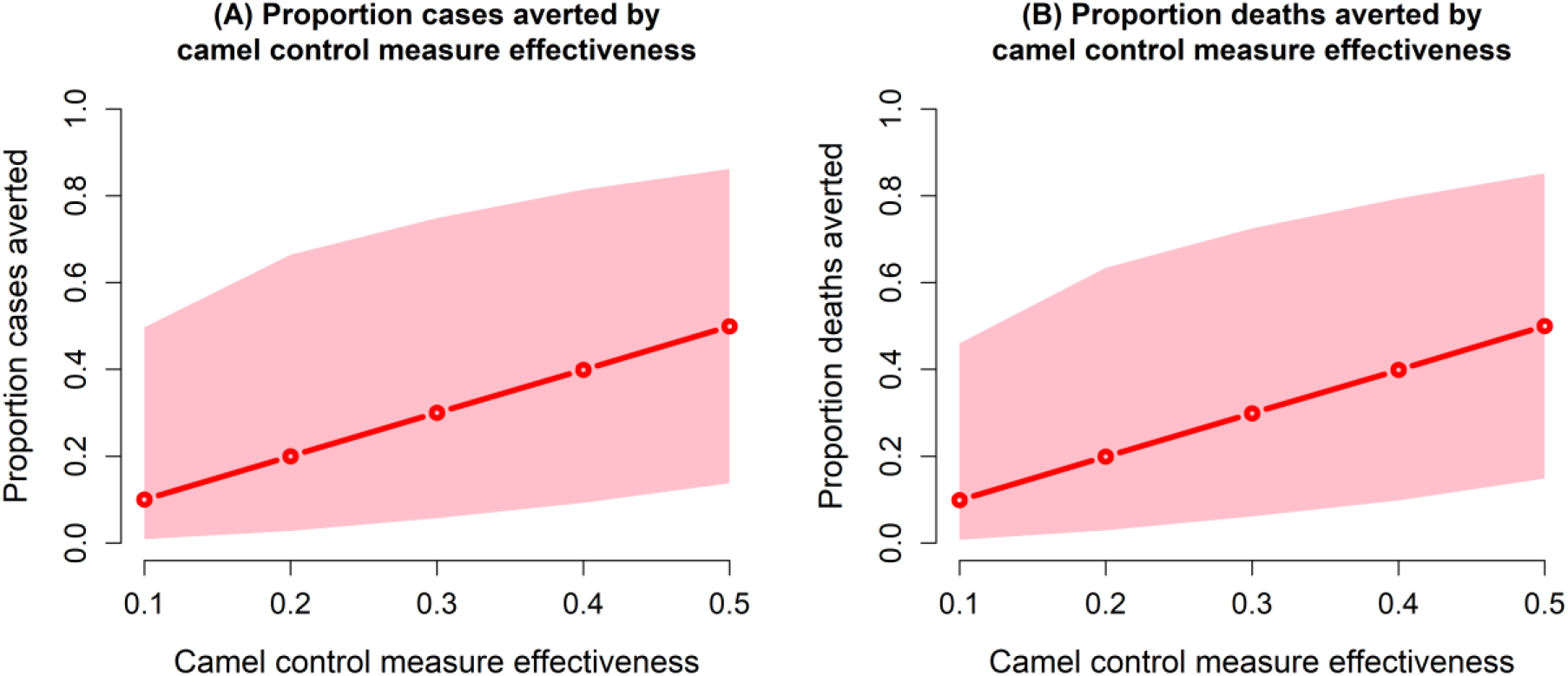
Proportions of cases (A) and deaths (B) averted with effectiveness of camel control measures. Mean proportion in red and 95% credible intervals shown in pink.

**Figure S2.**
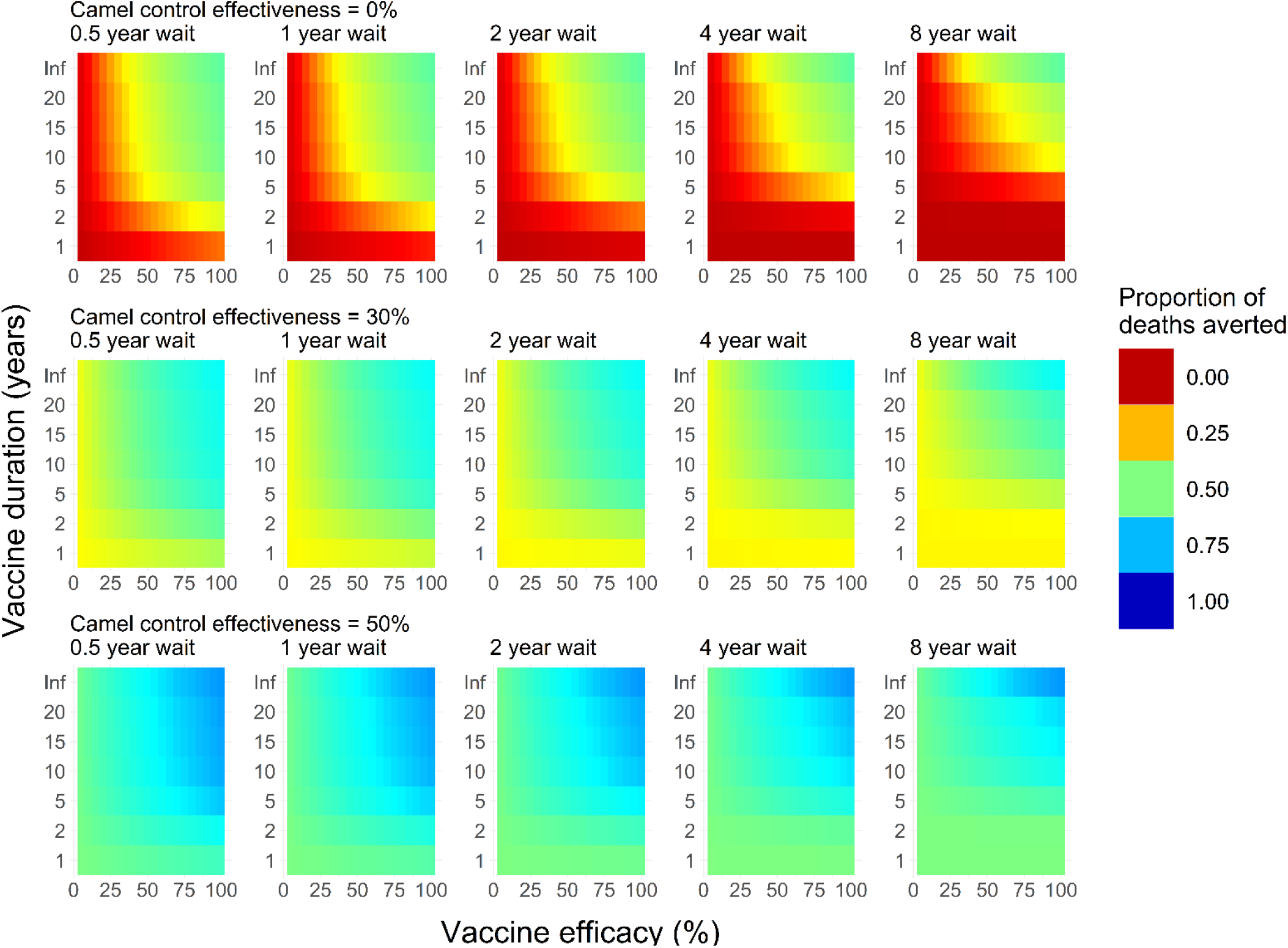
As per Figure 4, but each plot shows the mean posterior estimate of the proportion of deaths averted.

**Figure S3.**
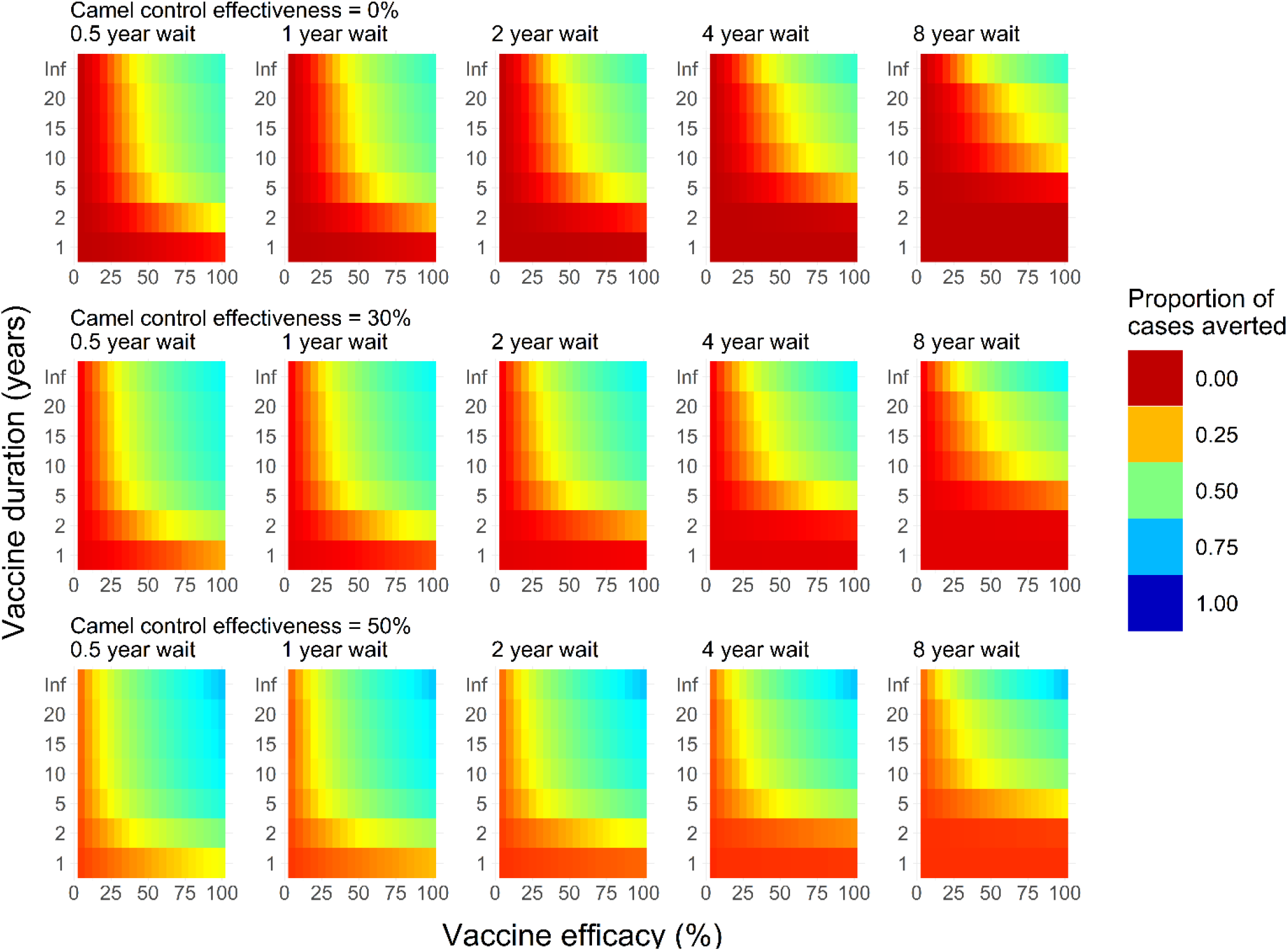
As per Figure 4, but showing lower credible intervals of the proportion of cases averted.

**Figure S4.**
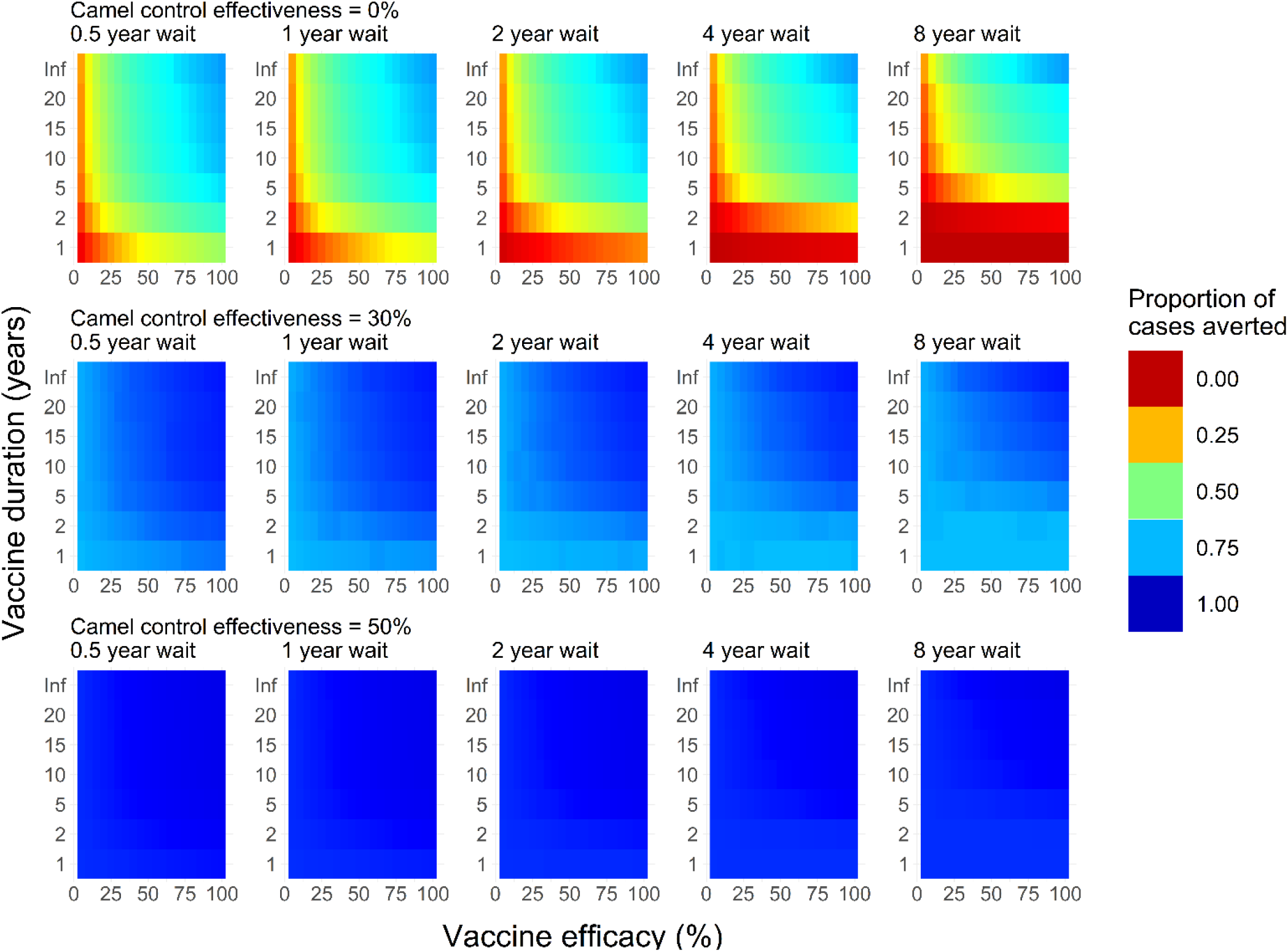
As per Figure 4, but showing upper credible intervals of the proportion of cases averted.

**Figure S5.**
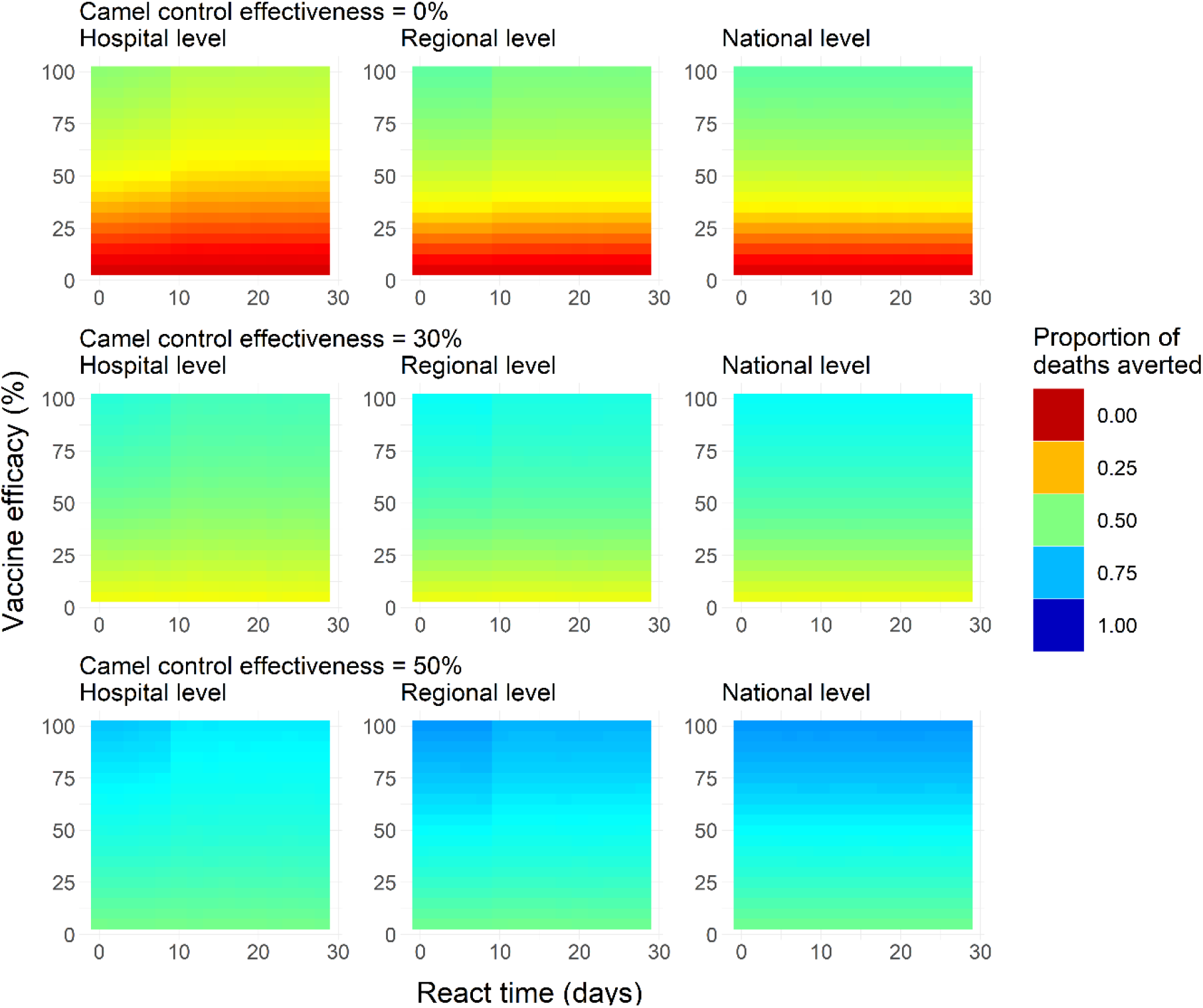
As per Figure 5, but showing mean posterior estimates of the proportion of deaths averted.

**Figure S6.**
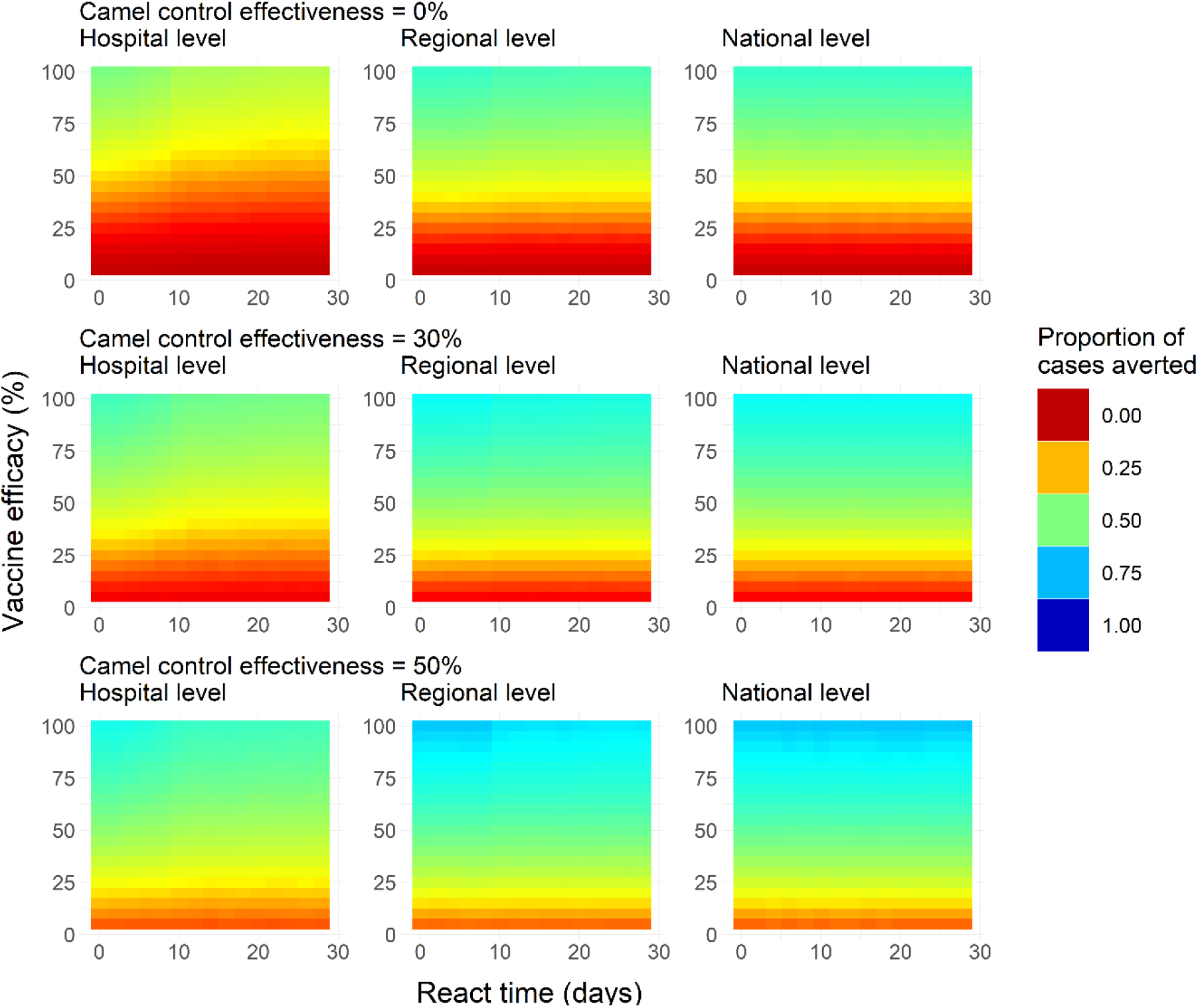
As per Figure 5, for lower credible intervals of proportion of cases averted as a function of efficacy, react time and reaction level.

**Figure S7.**
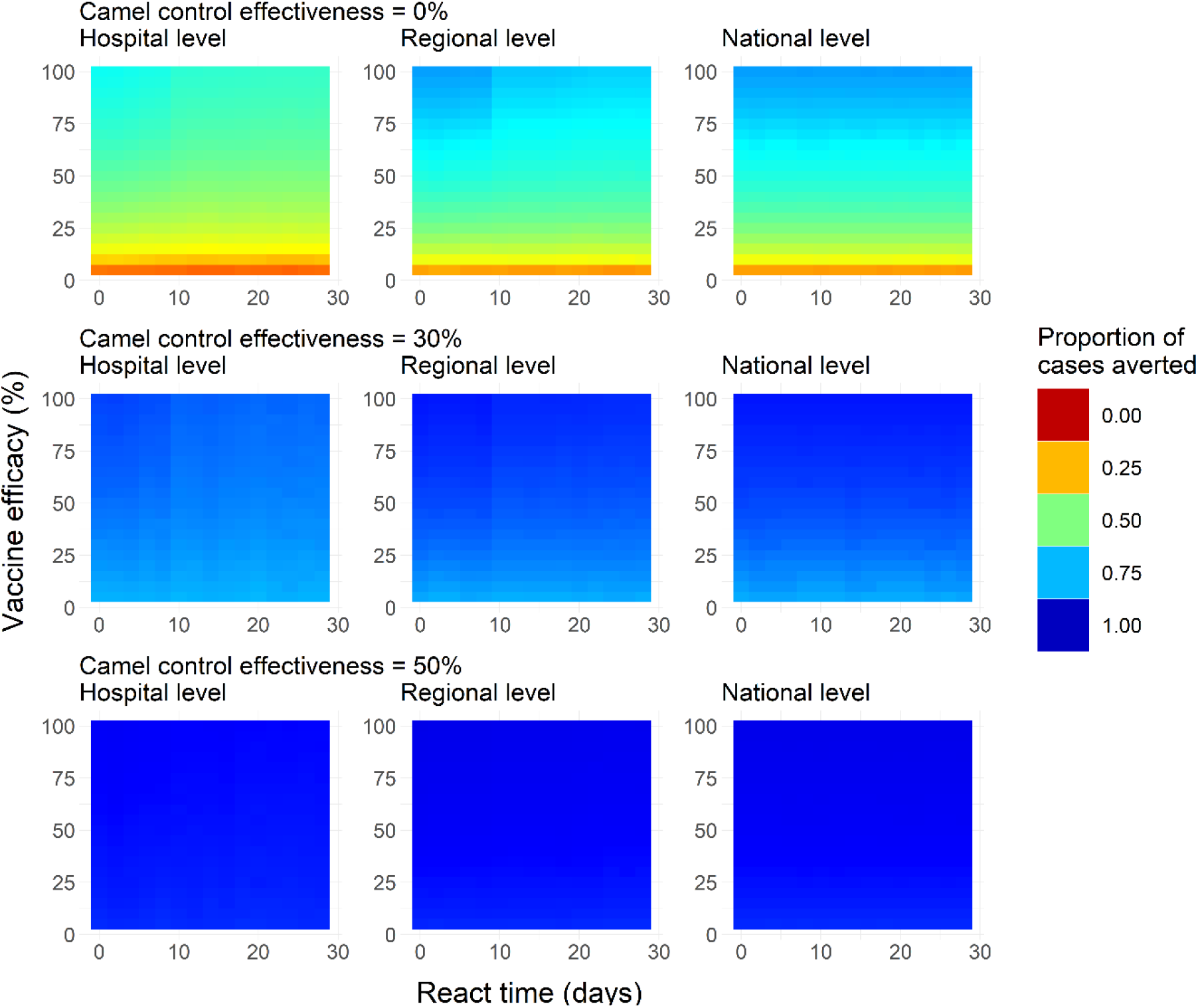
As per Figure 5, for upper credible intervals of proportion of cases averted as a function of efficacy, react time and reaction level.

**Figure S8.**
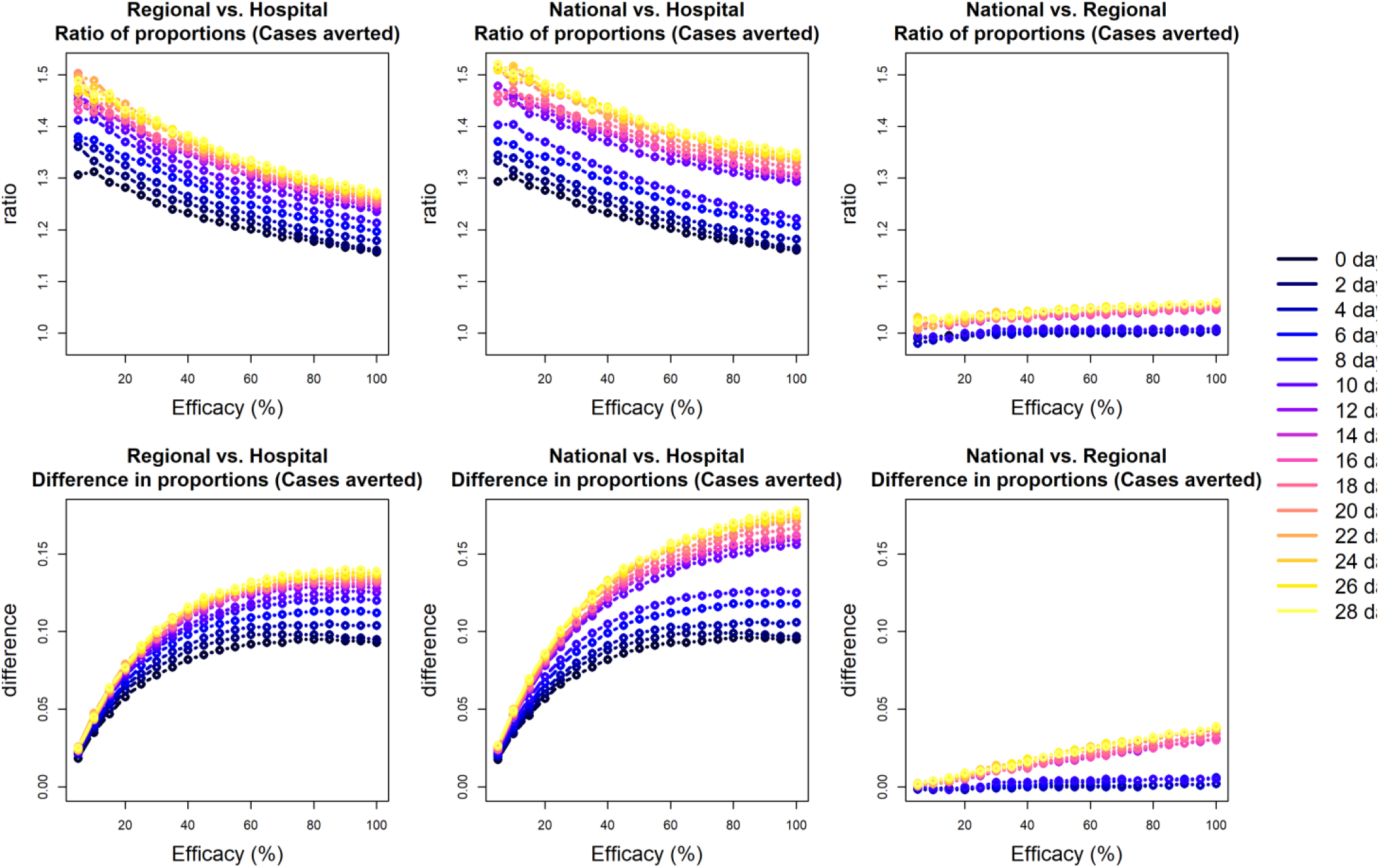
Comparison of reactive campaign levels. Top and bottom rows respectively show the ratios of and differences between the proportions of cases averted, by react time and efficacy. Left, middle and right columns show: i) regional vs. hospital; ii) national vs. hospital; iii) national vs. regional.

**Figure S9.**
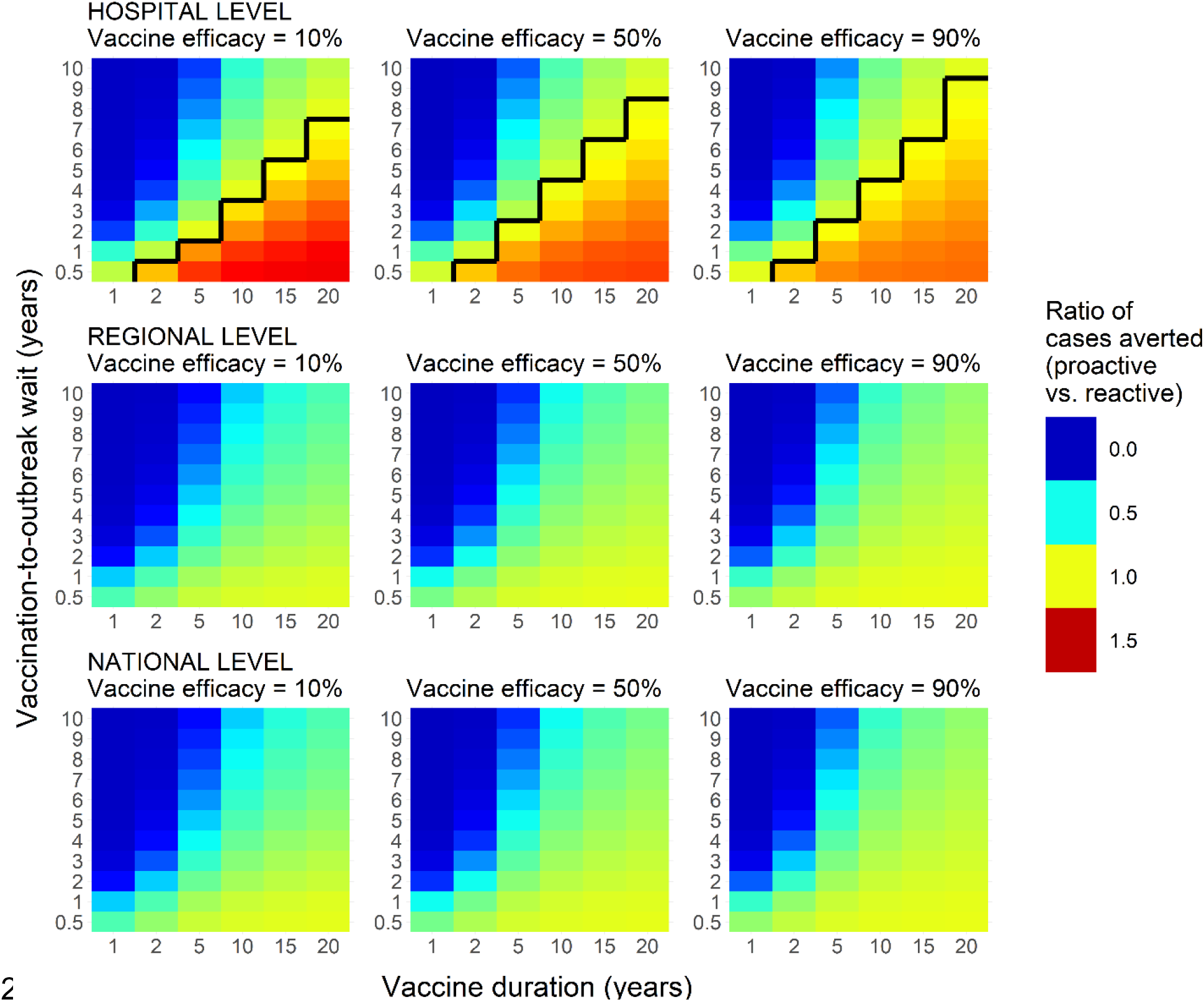
As per Figure 6, but assuming an 8-day react time.

**Figure S10.**
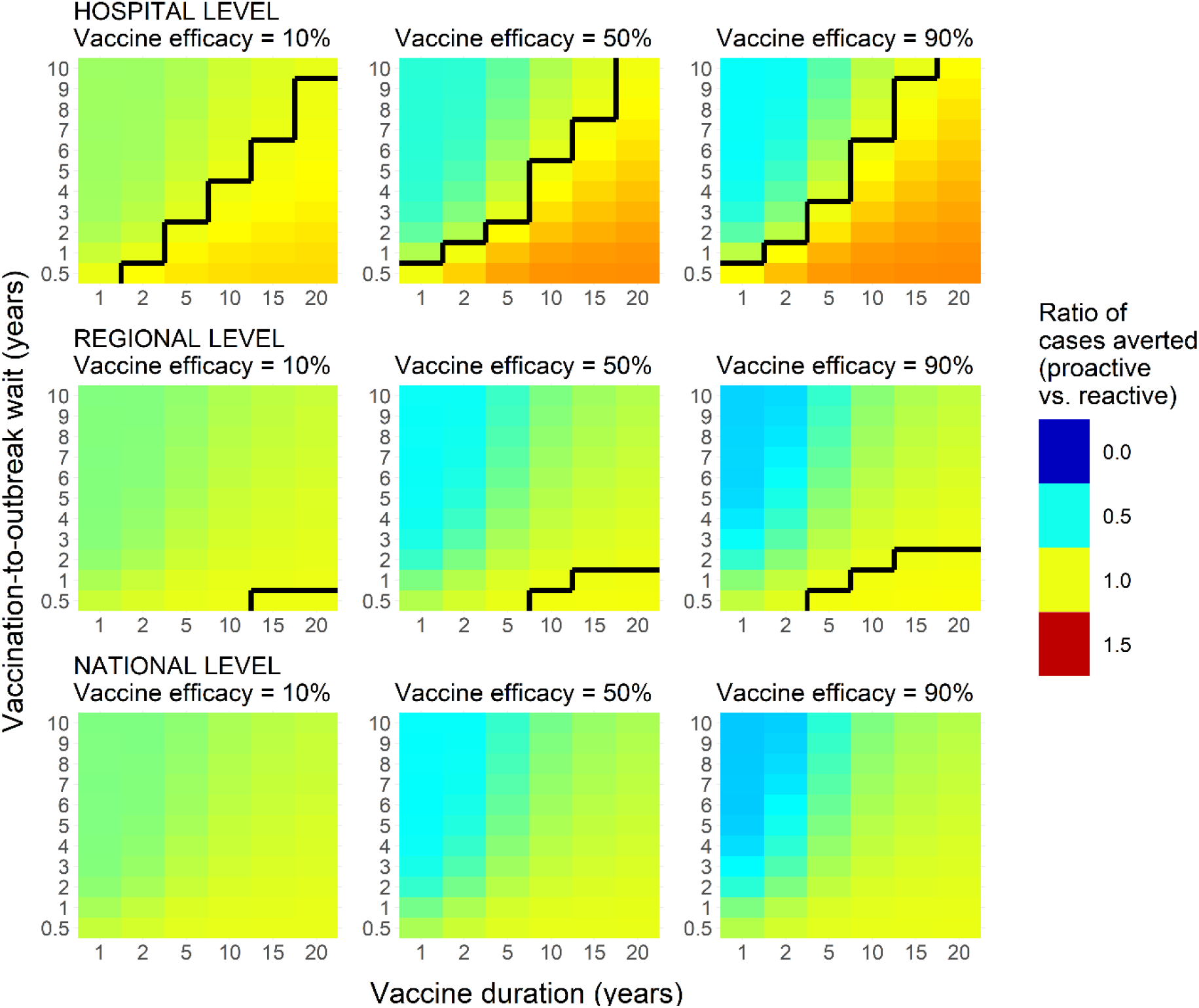
As per Figure 6, but assuming 30% effective camel control measures in tandem with both proactive and reactive campaigns.

**Figure S11.**
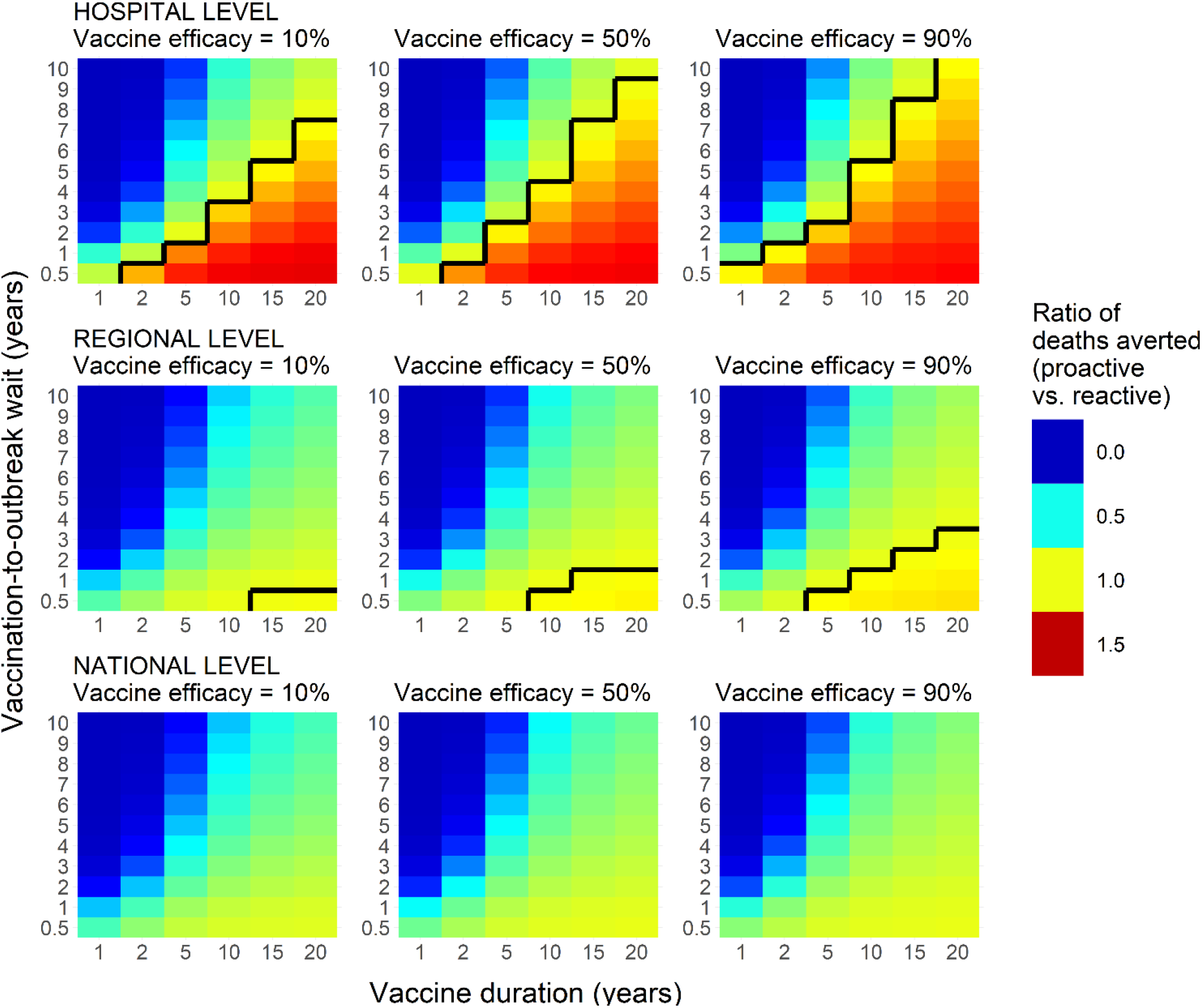
As per Figure 6, but showing ratio of deaths averted, as opposed to cases.

**Figure S12.**
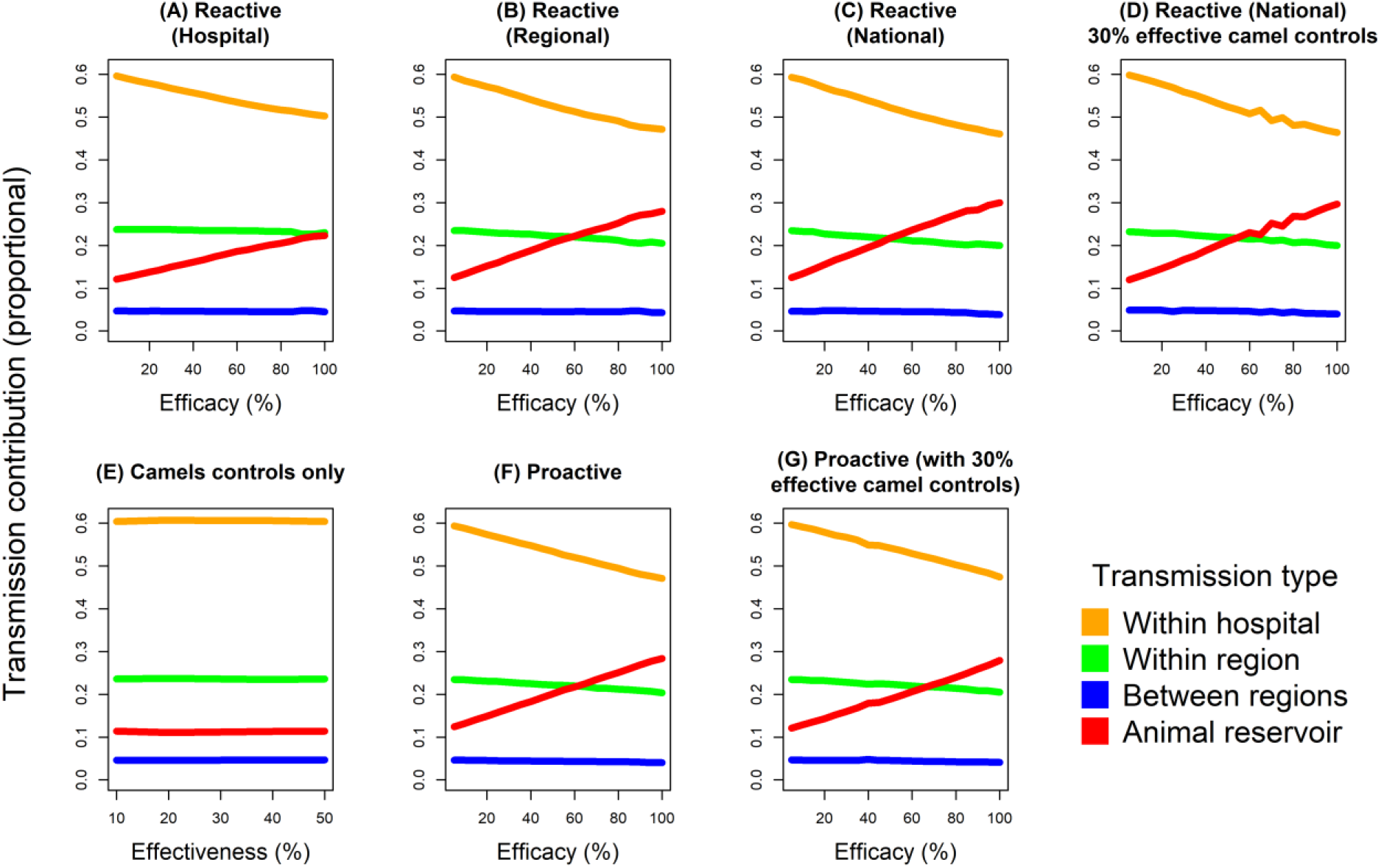
Change in relative contributions of transmission types with efficacy. Each plot shows the change in the proportion of overall cases numbers due to i) nosocomial; ii) regional; iii) national transmission and iv) the animal reservoir. Seven example vaccination campaign scenarios are shown: **(A)** reactive at hospital level; **(B)** reactive at regional level; **(C)** reactive at national level; **(D)** reactive at national level with 30% effective controls targeting camels; **(E)** Camel controls only (note that x-axis here shows effectiveness of camel control measures, not vaccine efficacy as per other plots); **(F)** proactive campaign without camel control measures and **(G)** proactive campaign with 30% effective camel control measures. Reactive campaigns assume 28-day react time. Proactive campaigns assume 5-year mean duration and 6 months wait until next outbreak.

**Figure S13.**
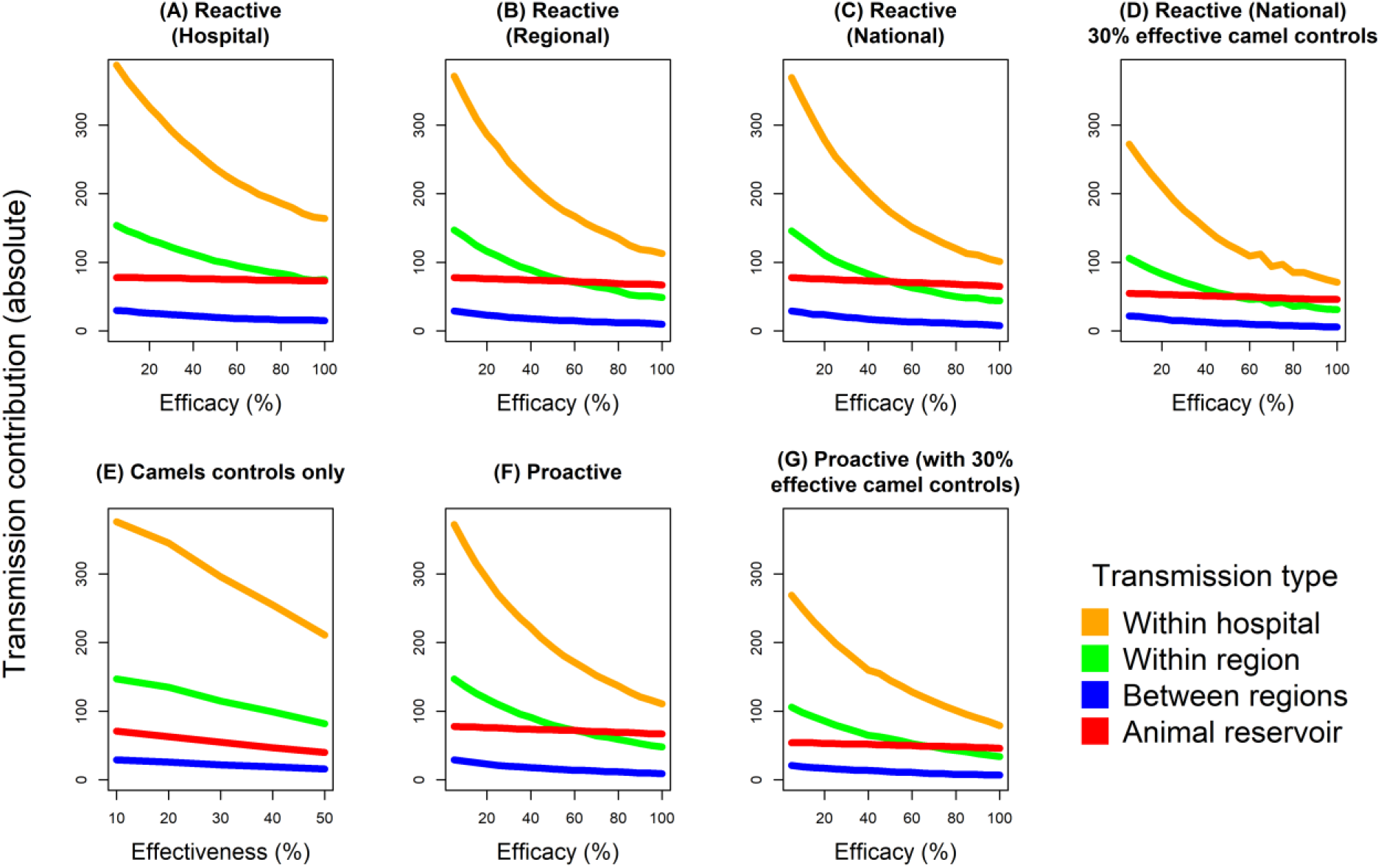
Change in absolute contributions of transmission types with efficacy. As per Figure S12 but showing absolute case numbers by transmission type and efficacy / effectiveness, not relative contributions.

**Figure S14.**
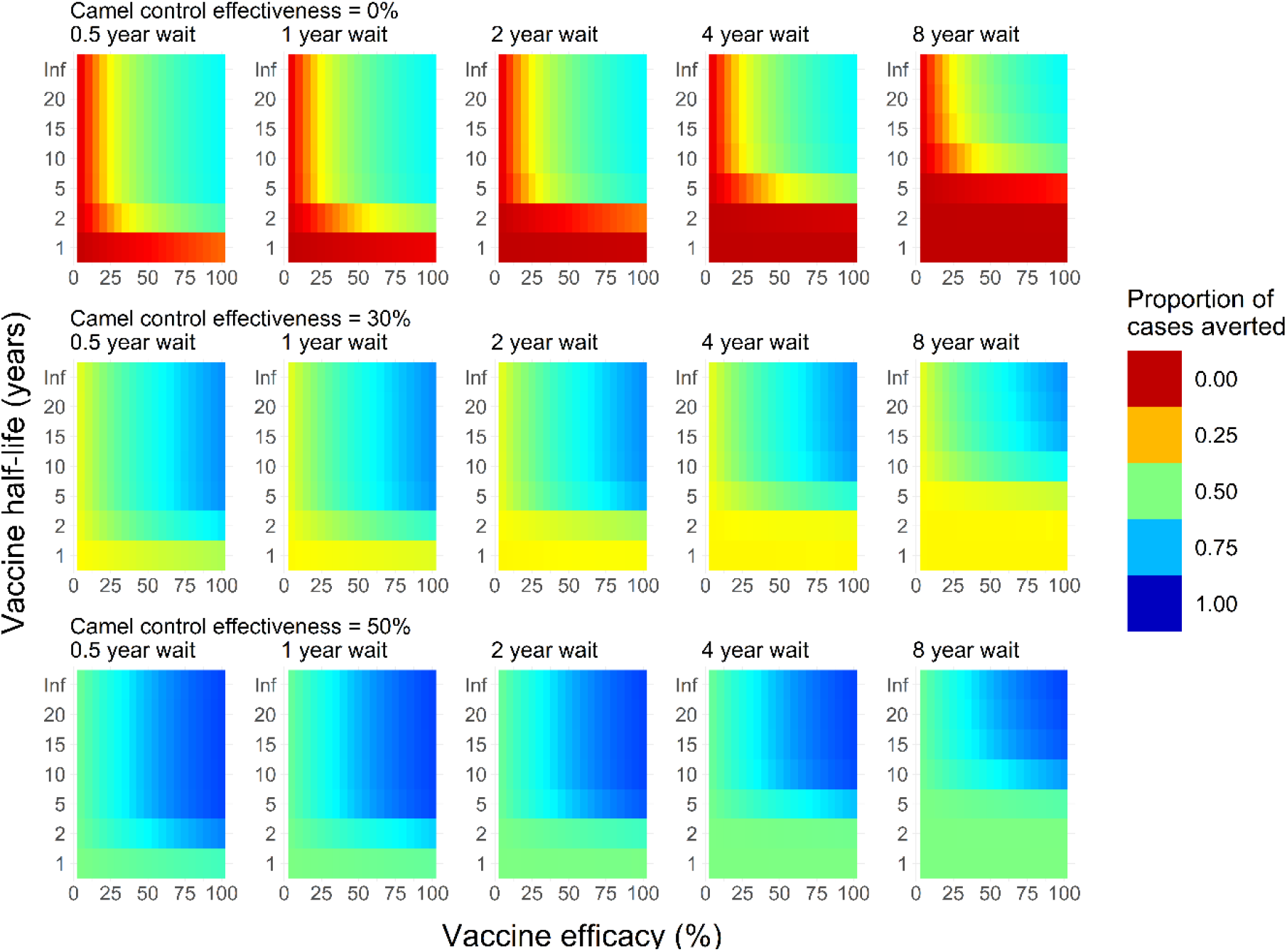
As per Figure 4, but showing mean posterior estimates of the proportion of cases averted in proactive campaigns assuming that vaccine efficacy wanes sigmoidally, and not exponentially as in our main analysis. Note that the y-axes denote vaccine efficacy half-life, as opposed to mean duration.

**Figure S15.**
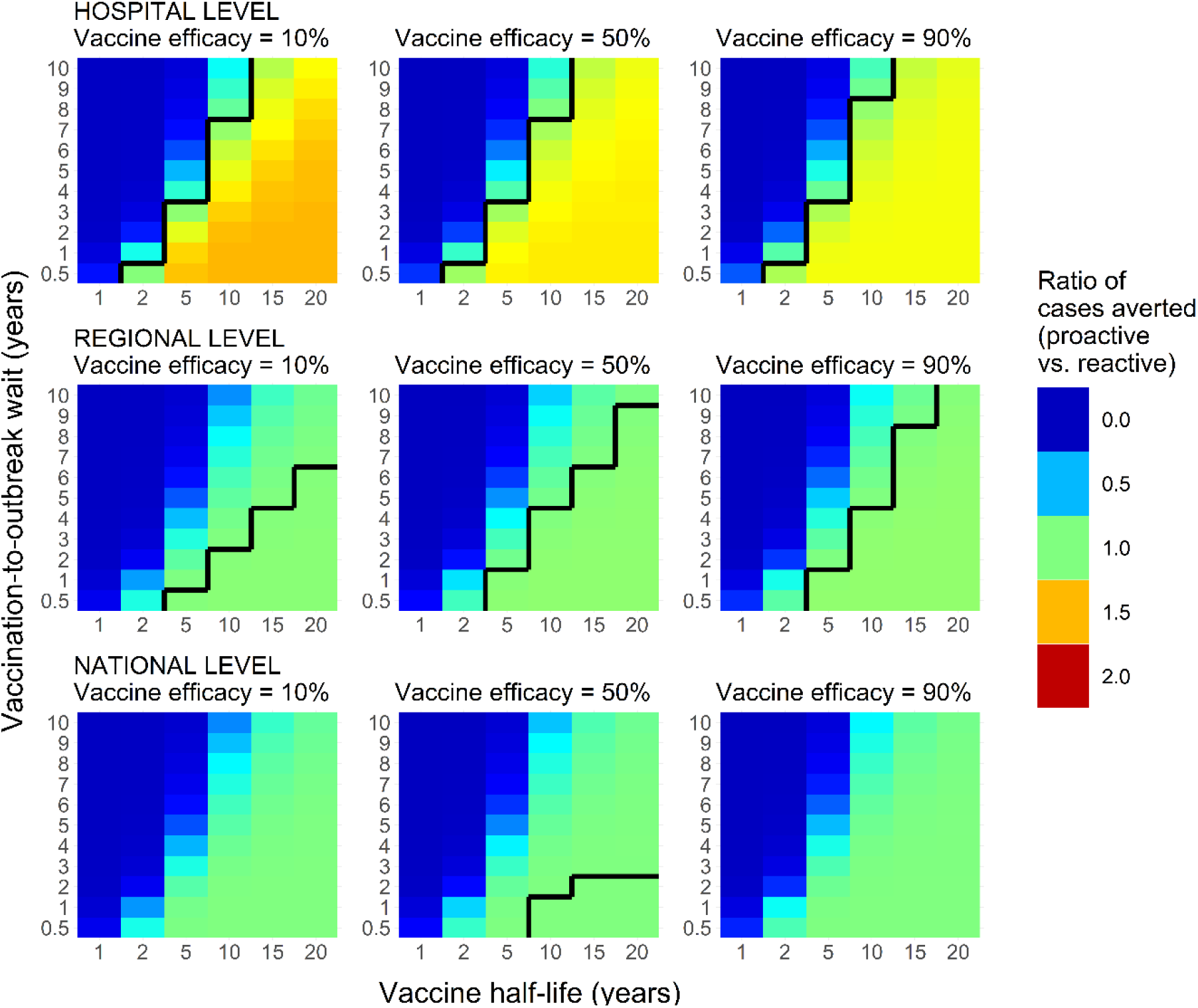
As per Figure 6, but where vaccine efficacy wanes sigmoidally, and not exponentially as in our main analysis. Note that the y-axes denote vaccine efficacy half-life, as opposed to mean duration.

**Figure S16.**
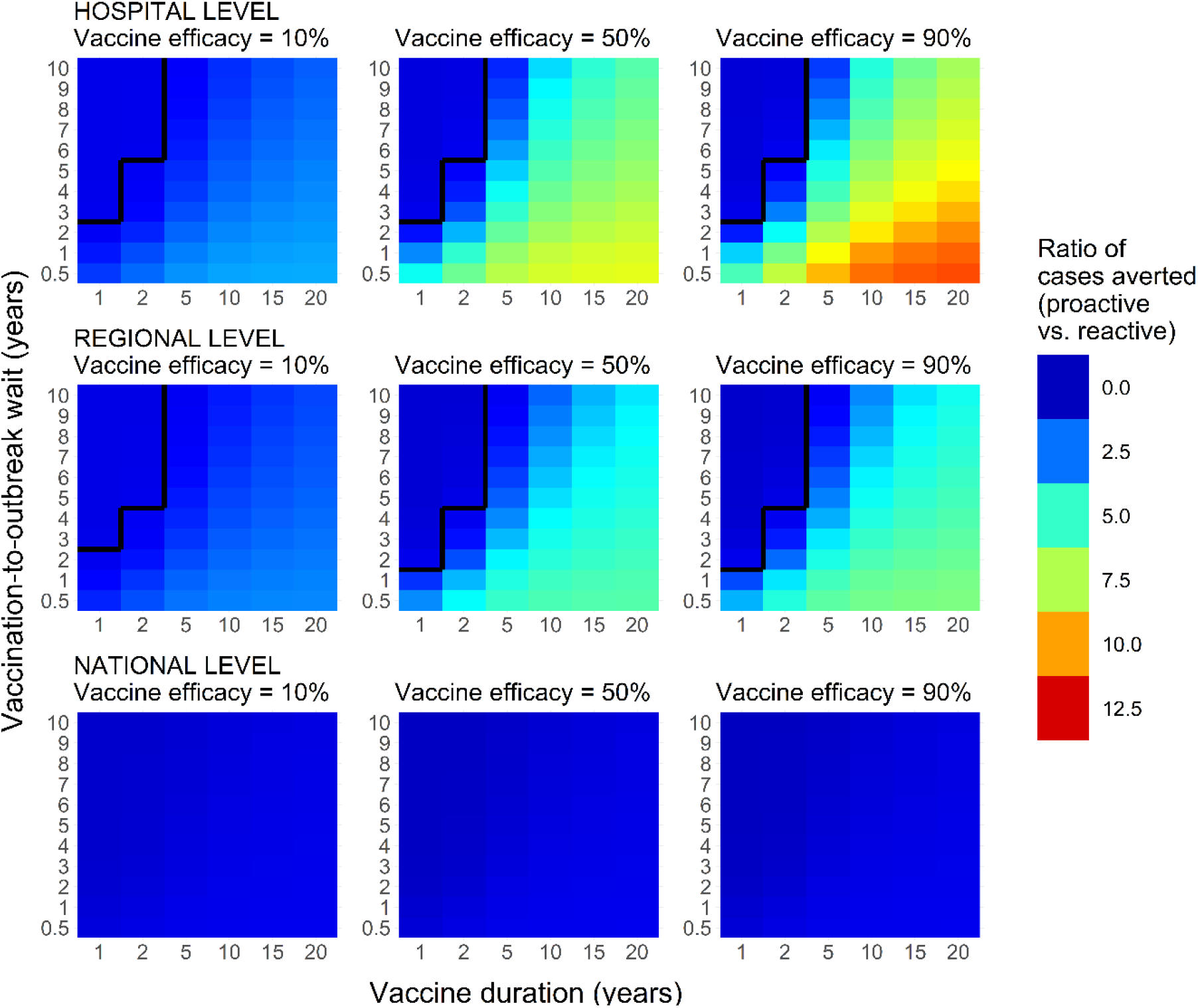
As per Figure 6, but showing mean posterior estimates of the proportion of cases averted if considering vaccination only during Jan-Jun 2013.

**Figure S17.**
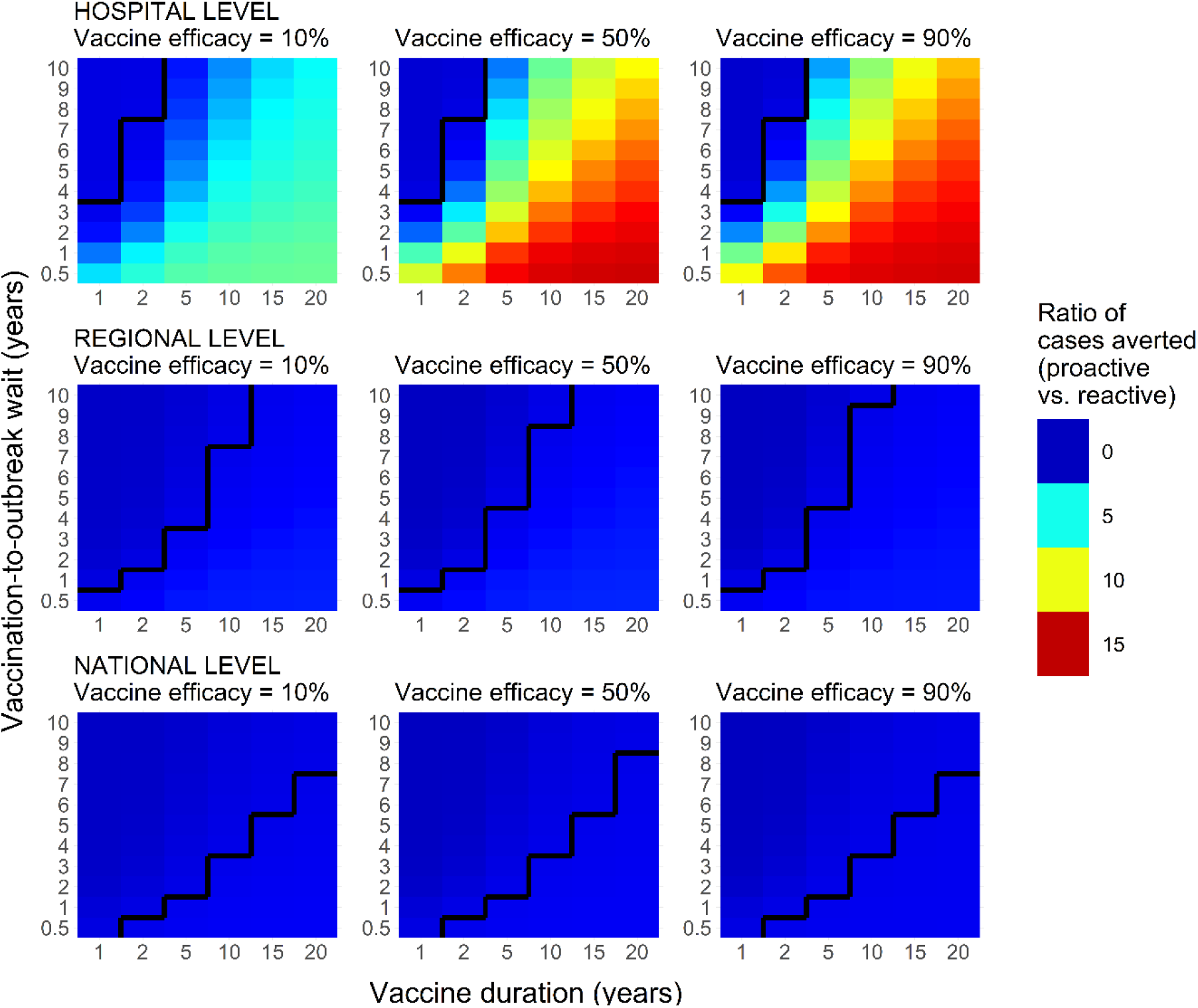
As per Figure 6, but showing mean posterior estimates of the proportion of cases averted if considering vaccination only during Jul-Dec 2013.

**Figure S18.**
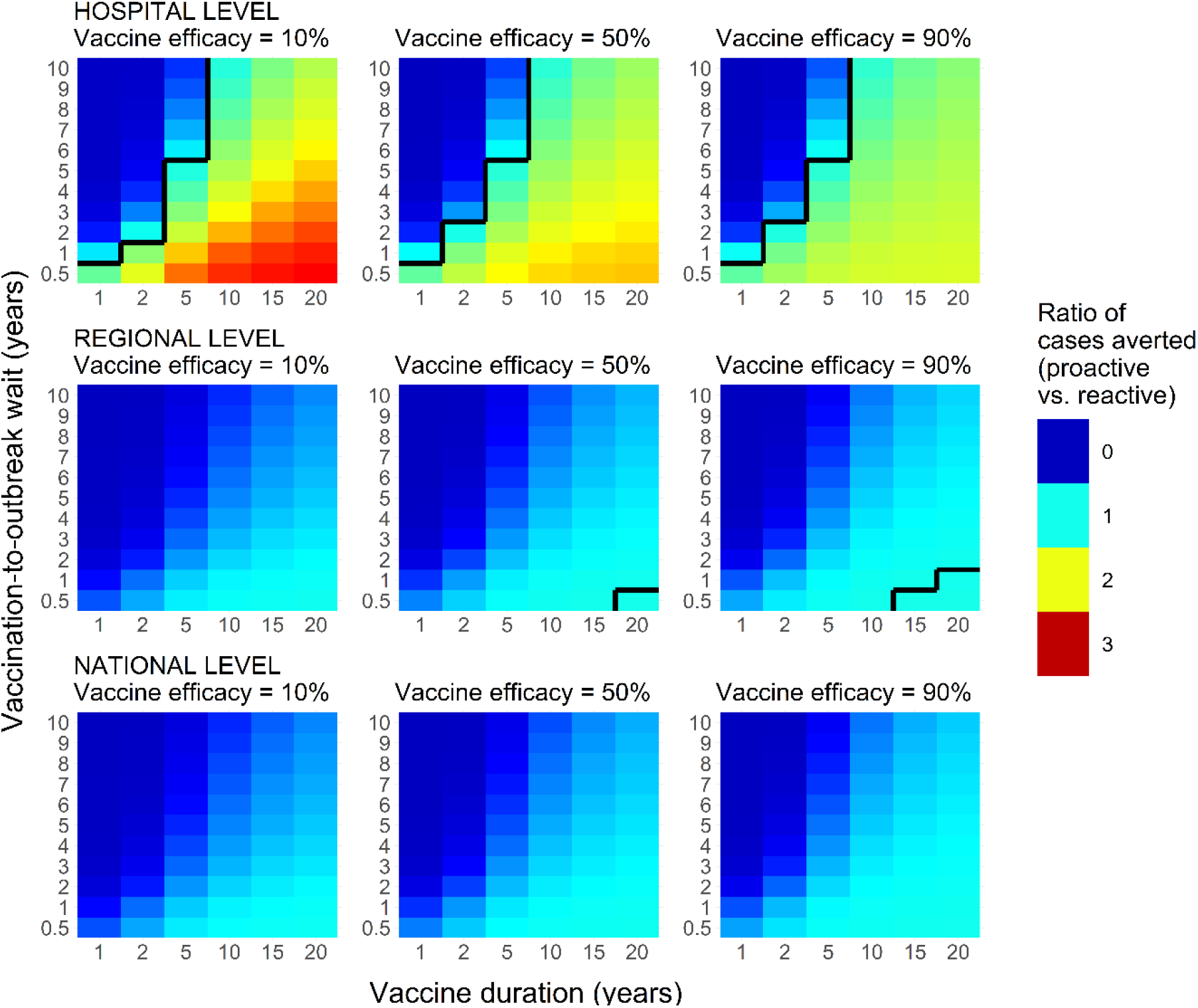
As per Figure 6, but showing mean posterior estimates of the proportion of cases averted if considering vaccination only during Jan-Jul 2014.

